# The Household Burden of Sleeping Sickness: Out-of-Pocket Costs for Diagnosis and Treatment

**DOI:** 10.64898/2026.05.05.26352145

**Authors:** R. Snijders, A. Fukinsia, P. Nganzobo, E. Miaka, E. Hasker, A. Mpanya, M. Antillon, F. Tediosi

## Abstract

**Purpose:** This study estimated out-of-pocket (OOP) expenses associated Human African Trypanosomiasis (HAT) care, in the Democratic Republic of the Congo (DRC) and explored how they influenced care-seeking behavior and participation in HAT control, aiming to inform effective and financially accessible elimination strategies.

**Methods:** A sequential mixed-methods study was conducted using 16 semi-structured interviews and 6 focus group discussions, followed by a structured survey of 444 recently tested participants across 6 health zones. Medical and non-medical expenditures were collected by health structure type and screening strategy (active vs. passive). Catastrophic health expenditure (CHE) was defined as OOP costs, excluding food, exceeding 10% or 25% of annual household income.

**Results:** Payments at health facilities, transport costs and long distances delayed care-seeking, particularly in passive screening (PS). Active screening (AS) was associated with minimal OOP, 93% of visits were cost-free, with a median OOP of 0.76 USD among those incurring costs. PS generated higher expenses, only 12% of PS visits were cost-free, with a median OOP of 9.08 USD among those with expenditures. Among confirmed cases, median OOP was lower through active (9.84 USD) than PS (24.23 USD). Nearly 90% of confirmed cases sold assets or borrowed money to cover expenses. CHE was uncommon under average household income(<4%), however 36% of passively detected cases exceeded the 10% threshold under minimum-wage income assumptions.

**Conclusion:** Despite free diagnosis and treatment, accessing HAT care in rural, low-resource foci in the DRC still imposes a substantial financial burden. Reaching elimination targets and ensuring equitable access will require minimizing indirect costs and logistical barriers to screening and diagnosis. As active screening declines, routine health systems assume greater surveillance responsibilities, reducing indirect costs and logistical these barriers will be critical to sustain coverage and maintain an effective and equitable HAT elimination strategy.

**What is already known on this topic:**

- Despite free-of-charge HAT screening and treatment in DRC, patients still incur substantial out-of-pocket costs (transport, food, informal/uncovered fees).
- These OOP costs deter timely care-seeking and screening participation, reducing or delaying treatment uptake and therefore threaten elimination efforts, yet no prior study had compared financial burden across active and passive screening strategies.

**What this study adds:**

- Contemporary, strategy-specific OOP estimates for gHAT in rural DRC, demonstrating that passive screening generates higher costs than active screening: 93% of active screening visits were cost-free versus only 12% in passive screening, with confirmed cases facing median OOP of $9.84 and $24.23 respectively.
- Evidence that costs escalate sharply with diagnostic delay — reaching a median of $141 for cases diagnosed at a fourth visit — and that nearly 90% of confirmed cases resorted to selling assets or borrowing to cover expenses, with 36% of passively detected cases exceeding catastrophic expenditure thresholds under minimum-wage income assumptions.
- A characterization of how financial barriers shape the full care-seeking pathway, from initial symptom response through diagnosis and treatment, including reliance on pharmacies and primary care facilities unable to diagnose gHAT, postponement of care in anticipation of mobile teams, and the deterrent effect of anticipated costs even when screening is nominally free.

**How this study might affect research, practice or policy:**

- Aims to make OOP costs (transport, food, uncovered fees) an agenda item in stakeholder meetings and community sensitization, including transparent communication of expected costs to reduce uncertainty as a barrier to care-seeking.
- Programme design should consider maintaining mobile active screening in rural, low-income foci where passive care pathway costs are highest, and target support measures — such as transport assistance and non-medical cost coverage during hospitalization — toward minimum-wage and asset-poor households most vulnerable to catastrophic expenditure.
- As elimination progresses and passive surveillance assumes greater importance, research is needed on strategies to improve uptake and reduce financial barriers, including transport vouchers, community-level incentives, and decentralization of diagnostic capacity to primary care facilities and pharmacies where patients already seek care.

## Introduction

Sleeping sickness or Human African Trypanosomiasis gambiense (gHAT) is a parasitic disease caused by the *Trypanosoma brucei gambiens*e, transmitted through tsetse fly bites and lethal when untreated.[1] During the colonial era, the disease devastated West and Central Africa, decimating entire villages.[2] Eventually, controlled through intensive case management and vector control, a resurgence in the 1970s led to nearly 40,000 annual cases by the late 1990s.[3] Subsequent interventions achieved elimination as a global public health problem (PHP) in 2020, defined by the World Health Organization (WHO) as fewer than 2,000 annual cases continent-wide and under 1 case per 10,000 inhabitants in at least 90% of endemic foci. [4, 5] The current goal is zero human transmission by 2030. In 2024, WHO reported 546 gHAT cases across 10 countries, with 60% in the Democratic Republic of the Congo (DRC-S1).[6-8]

gHAT control combines blanket population screening in endemic villages (active screening, AS), testing of self-presenting patients in fixed health facilities (passive screening, PS), and tsetse control in certain regions. Diagnosis begins with serological screening, followed by more complex laboratory tests if positive. Treatment evolved from stage-specific regimens requiring a lumbar puncture to determine the disease stage - safe and effective pentamidine injections for stage 1 and arsenic-based melarsoprol for stage 2, associated with severe adverse events [1] - to nifurtimox-eflornithine combination therapy (NECT) for second-stage disease from 2009.[9] Since 2020, Fexinidazole, the first non-stage specific all-oral treatment effective against both stages, and no longer requiring staging by lumbar puncture, has been the recommended first-line therapy except for severe stage 2. Early 2026, the European Medicines Agency approved Acoziborole as a single dose treatment for both stages. [10, 11]

Although screening and treatment are officially free, patients face indirect and uncovered direct costs—transportation, food, and non-HAT medical expenses—limiting access to care. [12-14] This is important given that in 2024, 74.6% of the Congolese population lived below the extreme poverty line ($2.15/day), [15] with two-thirds lacking basic needs, particularly in rural and conflict-affected areas.[16] Prior gHAT out-of-pocket (OOP) expenses studies identified their importance for health-seeking behavior. Gouteux (1985) in the Republic of Congo, and Robays (2007) in the DRC described how OOP contributed to lower participation rates in AS.[12, 13] Lutumba et al. (2007) estimated the economic burden at $164 per case, representing 43% of a rural household’s annual income, though only 5% was due to direct medical expenses.[14] However, these studies focused on AS through mobile teams under melarsoprol treatment era, whereas PS now accounts for 40%-50% of cases identified, and safter treatments combined with a decreasing prevalence might have altered care-seeking dynamics. [17]

As WHO targets zero gHAT transmission by 2030 under the Neglected Tropical Diseases (NTD) Roadmap, understanding economic barriers under current screening and treatment strategies becomes increasingly critical.[18, 19] This study assesses the financial burden of gHAT on individuals screened through active and passive screening, and to examine how costs affect care-seeking and participation, to inform more effective and equitable gHAT strategies.

## Materials and Methods

### Research setting

The study was conducted in Kwilu province, western DRC (78,441 km^2^) with five territories and twenty-four health zones. Each health zone serves around 100,000 to 200,000 people through a reference hospital and peripheral health centers.[20-23] Kwilu is among the most economically disadvantaged provinces in the DRC: more than half the population lives in severe poverty, with over 86% lack reliable access to education, healthcare, clean water, or adequate housing.[16] In 2019, Kwilu reported the highest number of gHAT cases nationally, with several health zones exceeding the PHP target of 1/10,000 inhabitants, as illustrated S2.

### Study Design

A sequential mixed-methods design was used. An exploratory qualitative phase identified key themes, OOP-related variables and community concerns regarding the burden of gHAT control and financial barriers to gHAT care. The subsequent quantitative phase measured medical and non-medical expenditures related to gHAT screening and treatment and examined how these costs influenced care-seeking behaviors.

### Study participants and sample selection strategy

Eligible participants were consenting adults (≥18 years) from high and low-endemic villages who participated in gHAT screening. The qualitative phase additionally included healthcare workers and community leaders. All participants provided written informed consent prior to enrolment.

The sampling database was constructed from the PNLTHA database combined with health-facility registers. For each village, random lists were generated of adults tested for gHAT in the preceding six months, stratified by screening strategy: mobile teams visits (AS) or attendance at fixed health facilities (PS).

From these lists, Participants were recruited using a purposive approach to ensure representation across five mutually exclusive screening outcome groups: serology negative, serology result unknown, serology performed with microscopy negative, serology positive with microscopy not done or unknown, and confirmed gHAT cases defined as parasitological confirmation by microscopy. Sampling across outcome groups and screening strategies was intentional to ensure heterogeneity in out-of-pocket costs across care pathways, as a purely random selection from the screened population would have yielded very few confirmed cases, underestimating the costs of gHAT treatment.

### Qualitative phase

Routine 2020 PNLTHA data were first reviewed to identify suitable endemic health zones (S2). Semi-structured thematic guides were developed around four domains: knowledge and perceptions of gHAT; care-seeking behaviors and therapeutic pathways; the influence of costs on choices; and factors shaping decisions to undergo gHAT screening.[24]

Between May and July 2021, semi-structured interviews (SSI) and focus group discussions (FGD) were organized until thematic saturation (thematic guides: S17 and S18). Interviews and FGD were conducted in Kikongo by an experienced anthropologist, audio-recorded, translated, and transcribed into French. Transcripts were imported and analyzed in NVivo 11 using a codebook based on the topic guides, expanded iteratively with inductive codes.

### Quantitative phase

Based on the qualitative results, a structured survey was developed in ODK (Open Data Kit), combined with standardized paper forms to capture all payments (Survey: S19).[25] In September 2021, six teams of trained local surveyors, accompanied by supervisors, contacted 449 people across six different health zones, covering two endemic health areas per health zone (S2).

Health structures were classified by screening strategy. Visits to traditional practitioners, pharmacies, primary care facilities^1^, private practices, hospitals, CDTCs (Centres for Diagnosis, Control and Treatment), and other fixed structures were grouped as passive screening, as these facilities rely on self-presenting patients. With the exception of hospitals, CDTCs, and a limited number of primary care facilities, most other facilities generally lack the capacity for gHAT testing. Visits by mobile team (vehicle or motorcycle) were classified as active screening, reflecting their role in community level screening.

All analyses were performed in R (version 2023.06.1).[26, 27]

Group differences in gHAT symptoms awareness and health facilities visited by screening strategy were assessed using independent two-sample t-tests, with Welch’s correction applied when variances were unequal.

#### Out-Of-Pocket expenses

Costs were recorded in Congolese Francs (CDF) and converted to US dollars using the September 2021 exchange rate ($1 = 1,982 CDF).[28] Total OOP expenditure per visit was defined as the sum of medical and non-medical costs:

OOP = Medical costs(C_Med_) + Non-medical costs(C_NonMed_) where

C_Med_ = Consultation + Laboratory tests + Other procedures + Medication + Hospitalization + Other medical costs

C_NonMed_ = Transport+ Food+ Other non-medical costs

For participants reporting multiple visits, OOP expenditures cost were summed across all visits, to capture financial burden. At a participant level, costs were classified as active or passive screening according to the structure that performed the first serological test.

The proportion of zero-cost visits was calculated for each screening strategy, with differences assessed using chi-square or Fisher’s exact tests. Predictors of incurring any OOP expenditure were examined using binomial logistic regression, with health structure type, travel time, transport mode, and screening outcome as predictors. Among participants reporting non-zero expenditure, a generalized linear model with gamma distribution and log link assessed factors associated with expenditure level. Model fit was evaluated using likelihood ratio tests. Results are presented as adjusted odds ratios with 95% confidence intervals. Given right-skewed expenditures, costs were summarized using medians, restricted to visits and participants reporting non-zero expenditures, plotted on log scale, and compared using Kruskal–Wallis tests.

#### Catastrophic health expenditure(CHE)

CHE occurs when OOP payments exceed a household’s capacity to pay. Given the absence of reliable household consumption or income data for the DRC, we used the Anker Living Reference value for Rural DRC of 2021 as an income proxy, as it was contemporary with our data collection period and most representative for rural settings. This reference estimates average monthly rural household consumption at 310,181 CDF ($158 monthly; $1,896 annually) and at 54,224 CDF ($28 monthly; $336 annually) for minimum wage households earning, based on a reference family of six with 1.77 full-time-equivalent workers.[29] CHE was defined using the budget-share approach: OOP expenditure (excluding food costs) exceeding 10% or 25% of estimated annual household income.[30]

### Ethics

The study was approved by the Institutional Review Board of the Institute of Tropical Medicine, Antwerp, Belgium (ref. 1493/21), and by the Ethics Committee of the Université Protestante au Congo, DRC(ref. CEUPC 0091).

## Results

### Qualitative phase

Data collection continued until thematic saturation, yielding 6 FGDs and 16 SSIs (details: S3). The analysis summarized emerging key themes which were used for the development of the quantitative survey.

#### Pathways to Care: gHAT Awareness and Symptom Response

Community members demonstrated varying levels of gHAT awareness. Symptoms like somnolence, weakness, and body pain were commonly mentioned, although knowledge on transmission and specific symptoms remained limited. gHAT was often described as life-threatening, with severity emphasized through accounts of death among undiagnosed or untreated community members.

When feeling unwell, participants reported first seeking pharmacy medication, visiting health facilities only after three or more days when symptoms worsened. Care was recommended more quickly for young children, perceived as more vulnerable. Participants reported using over-the-counter medications like paracetamol without diagnosis. Health centers or hospitals were described as preferred for persistent or worsening symptoms, though several identified barriers limited the feasibility of this option.

#### gHAT screening, diagnosis and treatment

Most participants were aware that screening could be conducted either by mobile teams visiting villages (AS) or at fixed health facilities (PS). Many participants had experience with AS, either door-to-door by motorbike teams or village-centered by truck-based teams, with some preferring door-to-door visits for greater privacy, shorter waiting times, and better fit with daily routines.

For PS, participants reported that local health centers were often unable to diagnose gHAT due to lack of appropriate equipment (microscope). Fixed diagnostic services were perceived as too few and typically located in distant hospitals.

#### Economic Burden and Implications of gHAT Screening and treatment

All participants noted that AS was free, some recalling that previously a card payment existed. They also mentioned that after AS campaigns, village chiefs sometimes encouraged voluntary food contributions as a gesture of thanks.

Regarding PS, respondents reported that while gHAT tests are free, health workers would refuse to attend to patients unless initial payments—such as registration or patient file fees—were made, leading to delays or avoidance of care, even when formal medical attention was recognized as necessary.

*“Money is a barrier to care because when you arrive at the health center, before they even touch you, you first have to buy a form, then come the tests. If you don’t have the money, you don’t go, which is why my mom keeps us at home even when we feel very sick”*(FGD with women, gHAT endemic village)

Hospital visits were considered more expensive, and less accessible, than primary care, requiring extensive preparation, including arranging travel, childcare, and time off work. Participants described families selling livestock or belongings to afford care.

*“I can give them(health care staff) a goat. …, among the nurses, whoever has money can pay for you and take the goat or the chicken, depending on what you have. …*., *my wife was sick, but I didn’t have any money, so we brought a piece of cloth we had at home so that they could treat my wife …”* (SSI, adult male, gHAT endemic village)

Fear of unaffordable care and potential refusal of services led participants to delay or defer care, with expected payments ranging from 2,500 CFA (≈$1.26) to nearly 200,000 CFA (≈$100). Participants also reported substantial non-medical costs including motorcycle travel, fuel, food, lodging, registration fees, supplementary diagnostics, as well as lost income.

*“I have five children. If one of them has sleeping sickness and I have to go to the Mission for treatment, …, I have to take the other four children with me. Otherwise, who would feed them? If I left them in the village, the worst could happen*.*”* (FGD with women, gHAT endemic village)

#### Barriers gHAT screening

Non-participation in AS was attributed less to unwillingness than to fear, stigma, and practical barriers. Fear of social exclusion, loss of income, or extended absence from home outweighed perceived benefits of early diagnosis. Work responsibilities at home or in the fields led to prioritizing daily obligations over seeking care. Fear of painful procedures, like lumbar puncture, being publicly identified as sick, and dietary restrictions also discouraged participation. Mistrust and misinformation, including confusion between gHAT and HIV screening, also contributed to hesitancy. One participant highlighted the influential role of the *chef du village*, the traditional village leader, in mobilizing community participation in AS. Limited engagement or trust by this authority could hinder screening uptake

Delays between screening and diagnosis were common, particularly when travel to distant confirmation centers was required. Although treatment was free once diagnosed, access was reported difficult due to poor roads, lack of transportation and long distances, sometimes requiring up to two days on foot. Some suspected cases preferred to wait for mobile teams rather than seeking health facility care, further delaying diagnosis and treatment.

*“… the child doesn’t know how to come here for treatment because the Mission is very far away; he can’t walk and has no means of transportation*.*”* (SSI, adult male former gHAT patient, gHAT endemic village)

Treatment at clinical trial sites was perceived as offering higher-quality care, better infrastructure, and free non-gHAT-related medical care and services, including food, leading some patients to delay treatment or travel further to access these trial sites.

*“now they send a car or motorcycle to take you to where they treat sleeping sickness. They will give you food, and everything is completely free, even your return to the village after treatment*.*”* (FGD with men, gHAT endemic village)

Despite the challenges mentioned, most participants expressed a willingness to engage with and recommend gHAT services—particularly when they were accessible, affordable, and clearly communicated.

### Quantitative phase

#### Sample description

Of 449 individuals contacted, 444(99%) were eligible for inclusion and included in the analysis (table 1). Five were excluded due to absence, lack of consent, or inability to recall gHAT screening. The included population was 57% female, with nearly 80% under 50 years old. Around half were screened through AS and half through PS. Given the purposive sampling strategy, confirmed and seropositive participants were deliberately oversampled relative to their population prevalence — which in high-endemic settings rarely exceeds 1–2% — to ensure adequate representation across the full cost spectrum. In the sample, 10% were confirmed gHAT cases, 21% had positive serology with negative microscopy, and 59% were seronegative. Approximately 10% had incomplete or unknown diagnostic out-comes.

**Table 1.** Description of study population.

| Population contacted | Active<br>(N=230) | Passive<br>(N=214) | Excluded from<br>analysis<br>(n=5) | Overall<br>(N=449) |
| --- | --- | --- | --- | --- |
| **Study Population** |  |  |  |  |
| Absent | 0 (0%) | 0 (0%) | 2 (40%) | 2 (0.4%) |
| No Informed consent | 0 (0%) | 0 (0%) | 1 (20%) | 1 (0.2%) |
| No recall gHAT screening | 0 (0%) | 0 (0%) | 2 (40%) | 2 (0.4%) |
| Eligible for inclusion | 230 (100%) | 214 (100%) | 5 (100%) | 444 (98.9%) |
| Population Included | Active<br>(N=230) | Passive<br>(N=214) |  | Overall<br>(N=444) |
| **Sex** |  |  |  |  |
| Male | 102 (44.3%) | 86 (40.2%) |  | 188 (42.3%) |
| Female | 128 (55.7%) | 128 (59.8%) |  | 256 (57.7%) |
| Missing | 0 (0%) | 0 (0%) |  | 0 (0%) |
| **Age Category** |  |  |  |  |
| Young Adults: 18–30 years | 106 (46.1%) | 91 (42.5%) |  | 197 (44.4%) |
| Middle-Aged Adults: 31–50 years | 71 (30.9%) | 82 (38.3%) |  | 153 (34.5%) |
| Older Adults: 51–70 years | 47 (20.4%) | 36 (16.8%) |  | 83 (18.7%) |
| Seniors: 71+ years | 6 (2.6%) | 5 (2.3%) |  | 11 (2.5%) |
| Missing | 0 (0%) | 0 (0%) |  | 0 (0%) |
| **Screening outcome** |  |  |  |  |
| Confirmed gHAT case | 18 (7.8%) | 28 (13.1%) |  | 46 (10.4%) |
| Serology: result unknown | 10 (4.3%) | 11 (5.1%) |  | 21 (4.7%) |
| Serology performed; Microscopy negative | 61 (26.5%) | 32 (15.0%) |  | 93 (20.9%) |
| Serology: negative | 128 (55.7%) | 134 (62.6%) |  | 262 (59.0%) |
| Serology: positive; Microscopy: not done/unknown | 13 (5.7%) | 9 (4.2%) |  | 22 (5.0%) |

#### Knowledge of gHAT and health-seeking behavior

Participants identified on average three gHAT characteristics (S4), most commonly headaches, sleep disorders, and fever, with no significant difference between AS and PS (t = −0.92, df = 442, p = 0.356). Awareness of community gHAT cases was higher among AS participants (88%) than PS (73%)(S5).

Overall, 60% of participants reported perceived gHAT symptoms prior to screening, more commonly in PS (83%) than AS (38%) (S6). Among symptomatic participants, 65% sought care before screening (AS: 55/88; PS: 119/178), typically first attending a primary care facility (45%) or pharmacy (41%). Hospitals were rarely a first choice (9%), with over 80% turning to specialized services (mobile teams or CDTC) only after exhausting other options (S7).

Across 444 participants, 800 visits were reported (S8-10). Mean visits were slightly higher in the AS (1.92) compared to PS (1.67; t = 2.34, df = 422.9, p = 0.02). Pharmacies and mobile teams were most accessible, with over 95% of visits requiring less than one hour on foot. Primary care facilities were also generally accessible (71% within one hour) mainly reached by motorcycle (47%), walking (30%), or biking (20%). Hospital visits were more burdensome, with nearly half requiring half to a full day of travel, primarily by motorcycle (49%) or on foot (31%).

Income loss was reported by 54% during screening (241/444), 36% during microscopy confirmation (53/146) and 89% (41/46) during treatment, occurring more frequently in PS(S11). Most participants relied on agriculture and crop sales as main source of income, followed by fishing, while around 30% combined activities, such as farming, fishing, small-scale livestock, or selling palm products. Health insurance was rare (9/444) and the one insured gHAT case still incurred costs.

#### Out-of-pocket costs

PS was associated with substantially higher OOP expenditures than AS, both in likelihood of incurring expenditures and magnitude(Figure 1 & S12). Among AS visit, 93% were cost-free reporting, median costs of visits incurring OOP expenditures were $0.62 for vehicle-based AS and $2.02 for motorcycle-based. By contrast, 88% of PS visits generated costs, with median OOP expenditures fro paying visits ranging from $2.52 at pharmacies to $33.68 at hospitals.

**Figure 1:**
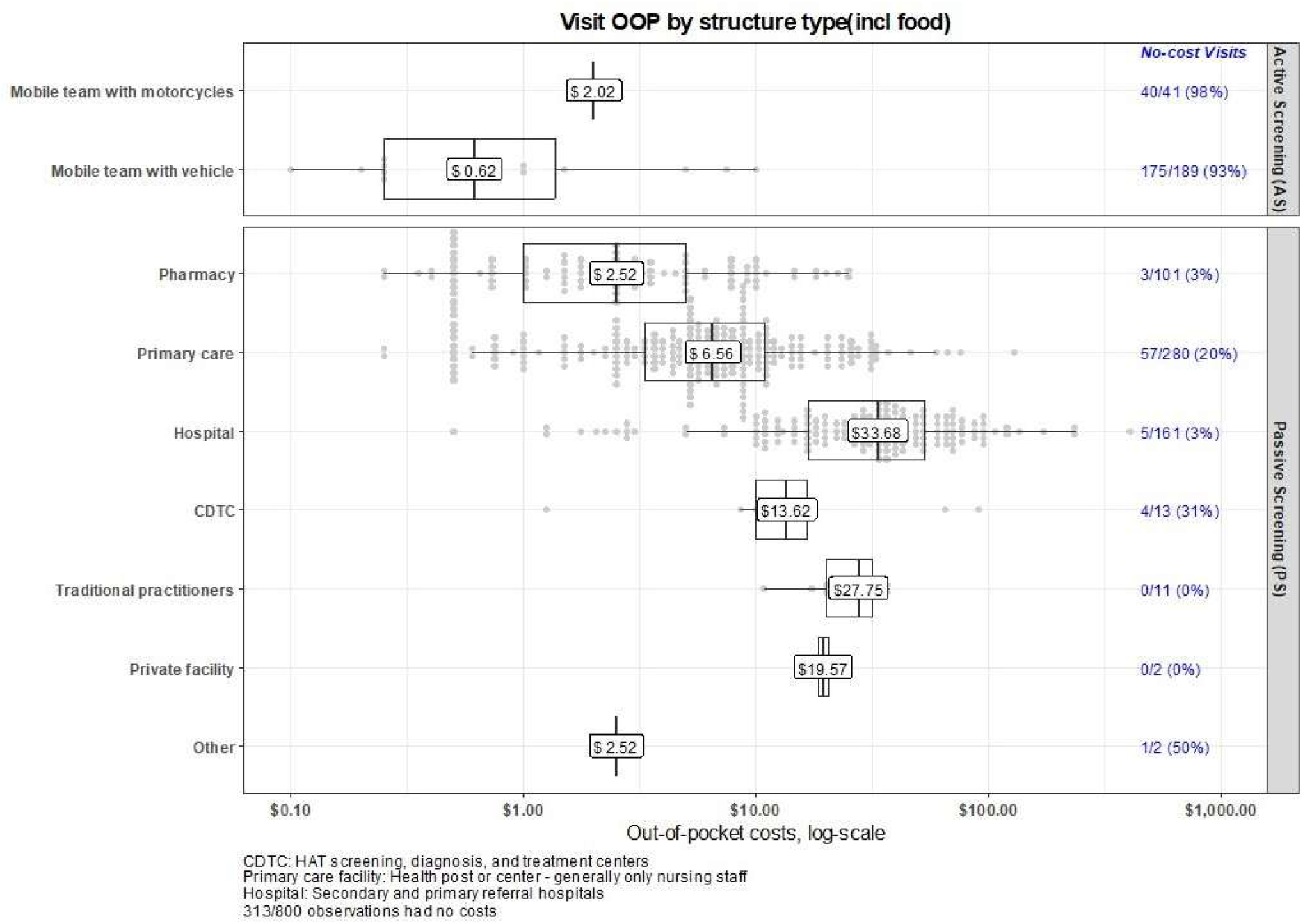
Boxplot of the median out-of-pocket expenses per visit by health structure type (visits with OOP > 0, including food costs) (n=800)

Regression analysis confirmed that health structure type was the strongest predictor of incurring OOP: visits to pharmacies, hospitals, and primary care facilities showed substantial higher odds of OOP expenditures (OR 42–442) compared to AS (S12). Travel time showed a borderline association (p = 0.057), while transport mode and screening outcome were not significant predictors. (S13). Across both strategies, approximately one-third of total OOP was attributed to medical care (medications, consumables, hospitalization), while another third went to food and transport (details S15-16).

Figure 2 shows OOP by health outcome and screening strategy across all 444 participants(S12). Across all outcome groups, 22%-77% of AS participants incurred no costs, compared to just 0-34% in PS. Among confirmed gHAT cases, median OOP was more than twice as high in PS ($24.23) than in AS ($9.84), and every PS-confirmed case incurred some cost, versus 22% cost-free in AS. Even seronegative participants faced a median OOP of $20.18 in PS, against $18.52 in AS among those reporting any costs, though far fewer AS participants incurred costs at all (51% cost-free vs. 11% in PS). OOP among AS participants with negative serological or microscopy results was largely driven by additional visits to fixed health structures outside of the mobile campaign than AS screening itself. In AS, 86% of participants requiring follow-up microscopy tests had it performed during the same mobile visit, limiting additional travel and follow-up (S9).

**Figure 2:**
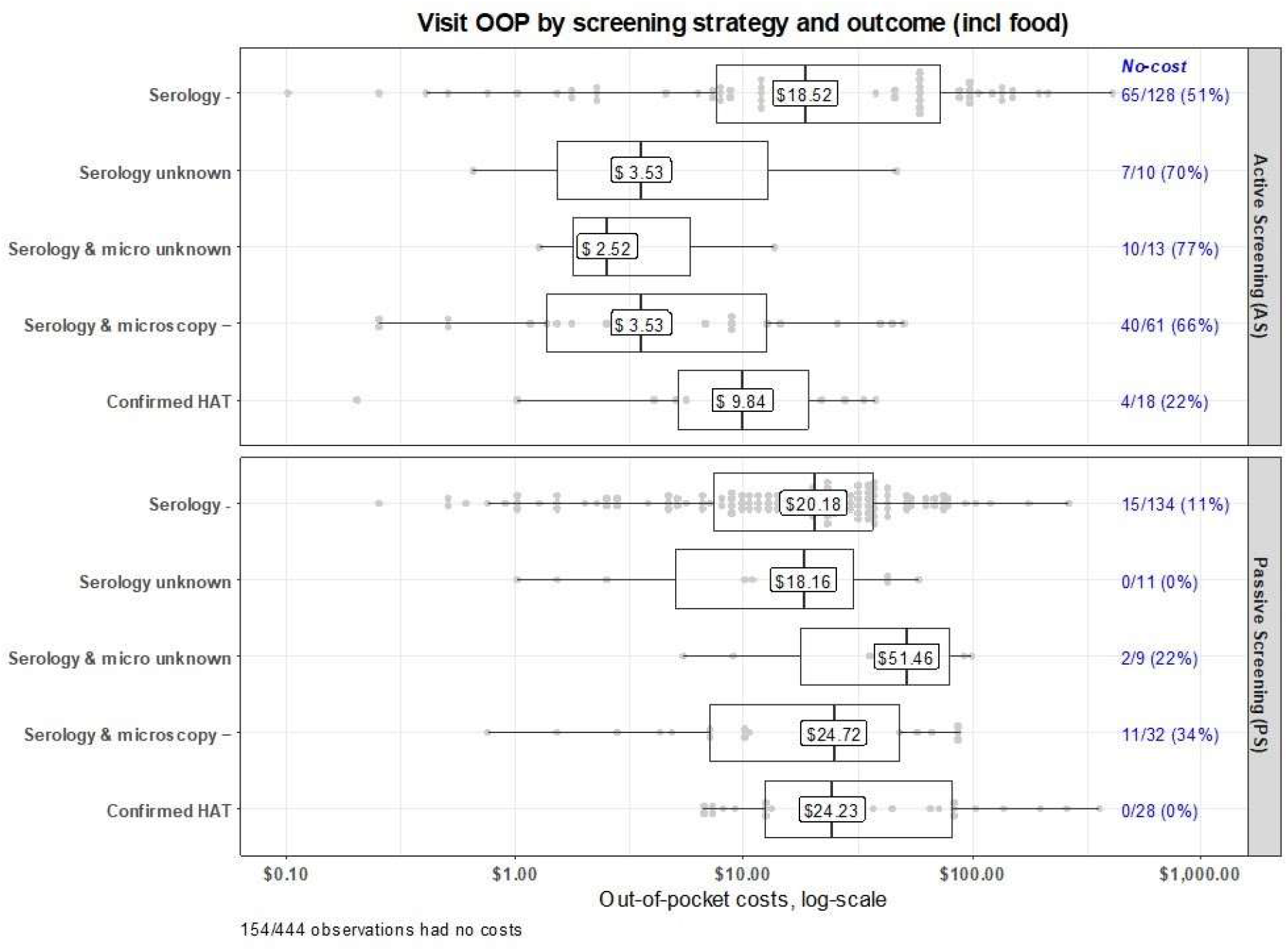
Boxplot of the median out-of-pocket expenses by screening outcome by screening strategy (visits with OOP > 0 – Including food costs) (n=444)

#### Confirmed cases

The 46 confirmed gHAT cases reported a median of two health structure visits (AS or PS) before diagnosis and treatment (S17). Nearly all (87%) reported symptoms prior to screening (AS: 14/18, 78%; AS: 26/28, 93%) and 44 out of 46 required hospitalization. While 40% began treatment immediately, 24% waited one to two weeks after arrival in the treatment facility. Most (76%) were hospitalized ten days or more, and nearly all (43/46) were accompanied by a family member.

Only four cases, diagnosed through AS, reported no OOP expenditures. AS-detected cases had a median OOP of $9.84 versus $24.53 for PS-detected cases (S12). Costs rose with diagnostic delay: median OOP was $8 for cases diagnosed at first visit, $21 at the second visit, $44 at the third visit, and $141 at a fourth visit.

Among confirmed cases medical costs accounted for 57% of average OOP, driven primarily by non-gHAT related medication (22%) and consultation fees (16%), while food (32%) and transport (10%), comprised the remainder (figure 3). To cope, nearly three-quarters resorted to selling assets, either alone (48%) or combined with other strategies (24%), with smaller proportions relying on family contributions (9%) or borrowing (6%).

**Figure 3:**
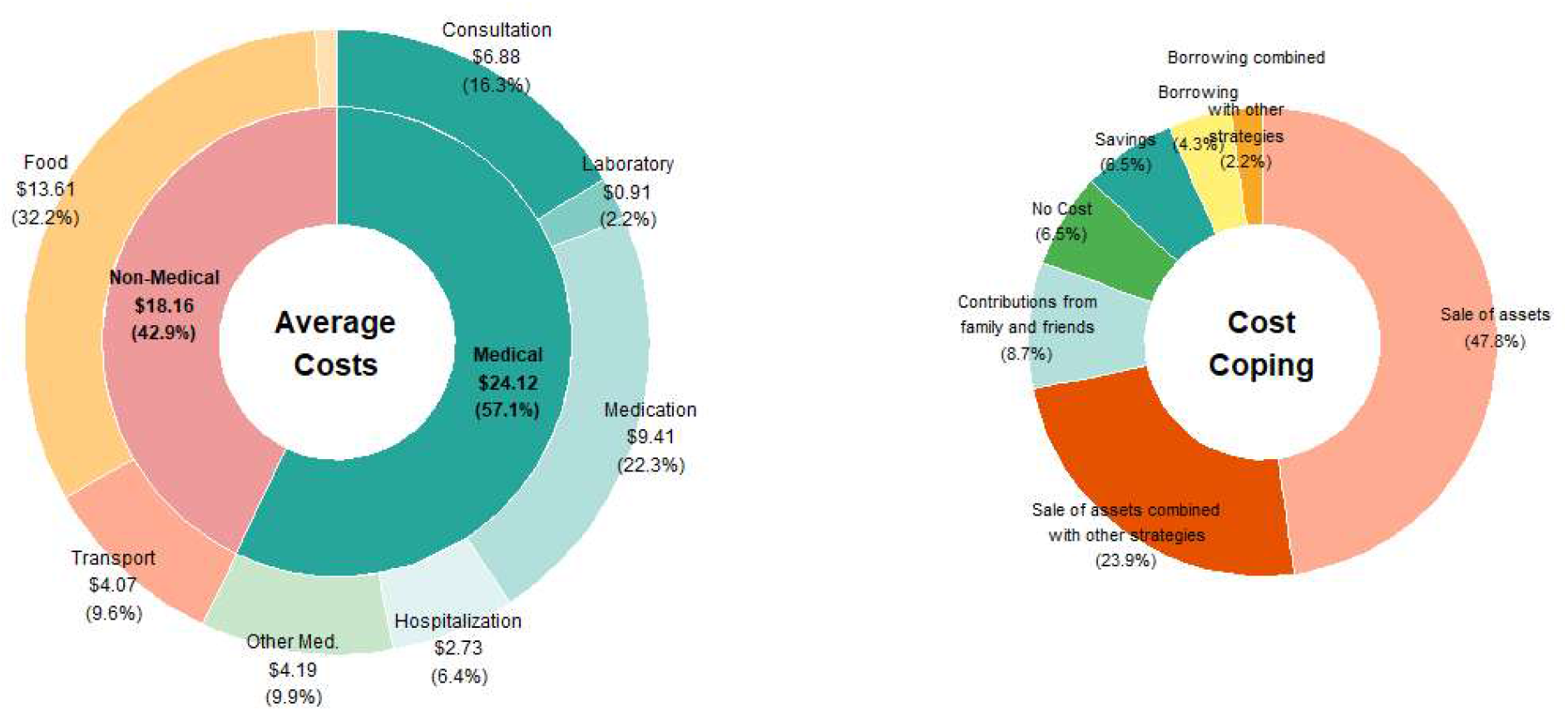
Out-of-pocket expenses ($) and financial coping strategies for confirmed gHAT cases.^2^

#### Catastrophic health expenditures

Based on the Anker Living Reference Value for rural DRC, CHE thresholds correspond to $190-474 annually for an average rural household, and $34-85 annually for a minimum wage household. Table 2 shows median OOP excluding food related expenses, stratified by screening strategy and screening outcome, and CHE prevalence defined as 10% and 25% of minimum and average wages.

**Table 2.**
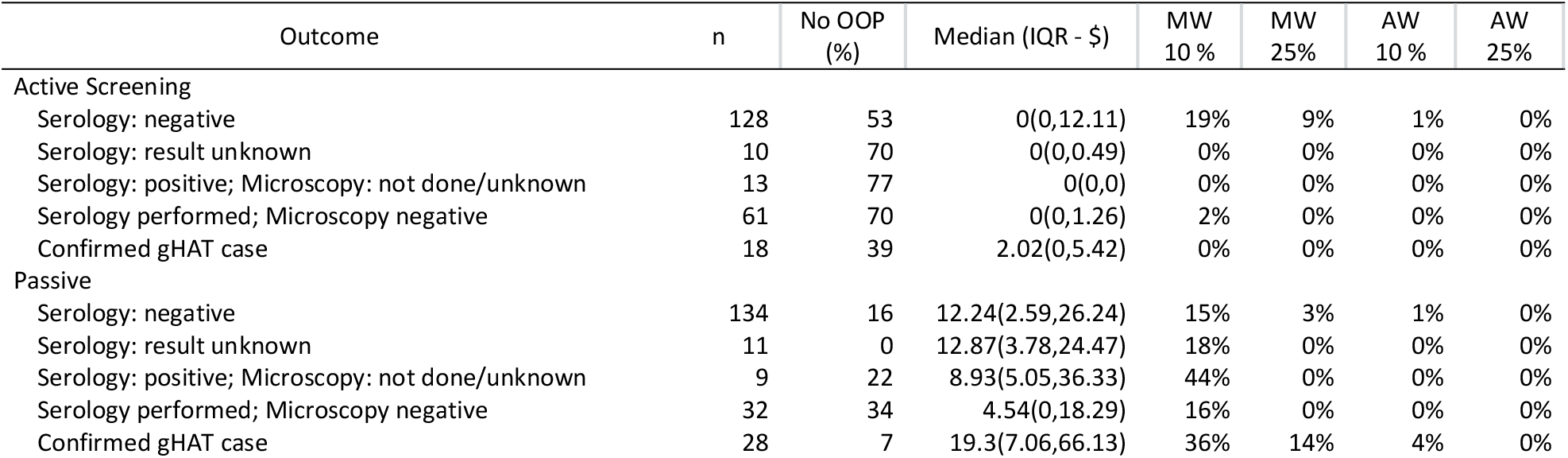
CHE prevalence by screening strategy and diagnostic outcome across income levels^3^.

| Outcome | n | No OOP (%) | Median (IQR - \$) | MW 10 % | MW 25 % | AW 10 % | AW 25 % |
| --- | --- | --- | --- | --- | --- | --- | --- |
| **Active Screening** |  |  |  |  |  |  |  |
| Serology: negative | 128 | 53 | 0(0,12.11) | 19% | 9% | 1% | 0% |
| Serology: result unknown | 10 | 70 | 0(0,0.49) | 0% | 0% | 0% | 0% |
| Serology: positive; Microscopy: not done/unknown | 13 | 77 | 0(0,0) | 0% | 0% | 0% | 0% |
| Serology performed; Microscopy negative | 61 | 70 | 0(0,1.26) | 2% | 0% | 0% | 0% |
| Confirmed gHAT case | 18 | 39 | 2.02(0,5.42) | 0% | 0% | 0% | 0% |
| **Passive** |  |  |  |  |  |  |  |
| Serology: negative | 134 | 16 | 12.24(2.59,26.24) | 15% | 3% | 1% | 0% |
| Serology: result unknown | 11 | 0 | 12.87(3.78,24.47) | 18% | 0% | 0% | 0% |
| Serology: positive; Microscopy: not done/unknown | 9 | 22 | 8.93(5.05,36.33) | 44% | 0% | 0% | 0% |
| Serology performed; Microscopy negative | 32 | 34 | 4.54(0,18.29) | 16% | 0% | 0% | 0% |
| Confirmed gHAT case | 28 | 7 | 19.3(7.06,66.13) | 36% | 14% | 4% | 0% |

<sup>2</sup> Part A: Average cost chart: Inner ring = sum of component averages; Part B are coping strategies for OOP diagnostic and treatment costs
<sup>3</sup> CHE = catastrophic health expenditure, defined as OOP exceeding specified percentage of annual household income; MW = minimum wage household (\$340/year); AW = average wage household (\$1,900/year). Thresholds: MW 10% = \$34; MW 25% = \$85; AW 10% = \$190; AW 25% = \$474.

AS participants carried the lowest financial burden, with the overall median OOP expenditure at $0.76, given that 39-77% of patients incurred no costs at all, and fewer than 1% exceeded CHE thresholds for average-wage households. Among seronegative AS participants, 19% exceeded the 10% minimum-wage threshold, likely reflecting costs incurred at fixed facilities visited outside the mobile campaign.

PS participants incurred higher OOP with only 7–34% avoiding OOP entirely, and confirmed gHAT cases had a median OOP of $19.30 [IQR: 7.06–66.13]). While OOP for PS participants rarely exceeded CHE thresholds for average-wage households (<4%), the burden is substantial for minimum-wage households: approximately one-fifth of passively screened participants exceeded the 10% annual income threshold, rising to 36% among confirmed cases. At the stricter 25% threshold for minimum-wage households, 14% of confirmed PS gHAT cases experienced catastrophic expenditure.

## Discussion

This study provides contemporary, screening strategy-specific estimates of OOP expenditure for gHAT screening, diagnosis and treatment in rural DRC, extending earlier work by comparing costs across AS and PS for the first time. [12-14] Although gHAT screening is officially free, participants still face substantial expenses. Median OOP remained low among in AS with 93% of visits cost-free, but was sub-stantially higher in PS ($20.51), particularly for confirmed cases ($24.52). While OOP rarely exceeded catastrophic health expenditure thresholds for average rural households, 36% of PS confirmed gHAT patients exceeded the 10% CHE minimum-wage income threshold. Overall, 89% of gHAT patients resorted to borrowing, selling assets, or seeking financial help. Only 9 of 444 participants had health insurance, and even the one insured gHAT patient incurred expenses—illustrating that gHAT endemic populations remain largely excluded from Universal Health Coverage (UHC) efforts. Which likely leads to delayed or foregone care, worse health outcomes, and a high risk of financial hardship due to out-of-pocket costs.

These findings carry implications for gHAT elimination strategies, which will increasingly depend on PS as AS scales back. The study revealed that community awareness of gHAT differed slightly by screening strategy (AS: 88% VS PS: 73%). As elimination progresses and cases become rarer, first-hand knowledge of cases will fade, reducing risk perception and community motivation to participate in screening, particularly in blanket population campaigns that rely on voluntary uptake.

The limitations of PS in rural low-income settings compound this challenge. The qualitative findings revealed that symptomatic individuals typically seek care first at low-cost, nearby options like pharmacies (41%) and primary care facilities (45%), which generally lack capacity for gHAT diagnosis beyond a symptom assessment. Hospitals capable of diagnosis were rarely selected as a first contact (9%) due to distance and cost such that over 45% of participants considering hospital visits only after exhausting other options. Some symptomatic patients chose to postpone care-seeking, waiting for mobile teams despite being aware of the risks. Specialized gHAT treatment centers were mentioned as a last resort, likely because they are embedded within regional hospitals and not clearly distinguished from the general health care system.

Geographic accessibility further constrains PS uptake. Over 75% of gHAT patients reached facilities within an hour on foot (≈5 km), suggesting that PS coverage is largely confined to populations living within a 5–10 km radius of diagnostic facilities, excluding at-risk individuals beyond this range. Simarro et al. estimated that 41% of Africa’s gHAT at-risk population lives within one hour of a screening facility and 71% within three hours.[31] Although WHO reported more PS sites in the DRC in 2024, these accessibility models typically assume the use of vehicles or motorcycles on main roads and tracks, thus overlooking key barriers such as transport affordability and availability in remote, low-resource settings.[5]

Several limitations of this study should be noted. First, income data was not collected from the participants due to technical complexity and social sensitivity. Instead, the Anker Living Reference Value was used as a reference. Second, the analysis did not include lost income, productivity, or caregiving costs, and likely underestimates the true economic burden. Additionally, exchange rate fluctuations ($1= 2,896 CDF; +46% in 2024 vs. July 2021) may have affected recorded OOP values, as internationally traded goods (drugs, fuel for transport) rose faster than local wages, therefore increasing the real burden.

We recommend actions focus on improving financial accessibility and patient awareness. OOP expenses, including transport, food, and uncovered medical fees, should feature prominently in stakeholder and community meetings to raise awareness of the real costs patients face. Maintaining mobile screening in rural, low-income foci, where PS-related costs are highest will remain essential for continued case detection and disease surveillance. Although AS poses a limited financial burden and is generally affordable even for the poorest households, fear of hidden costs may still discourage participation. Clear, standardized communication on the financial implications of screening and treatment, as well as available financial or logistical support, could reduce misconceptions and encourage earlier care-seeking.

However, as gHAT approaches elimination, maintaining stand-alone mobile screening may become unsustainable, and the declining perceived risk of the disease is likely to reduce community participation as other health and livelihood priorities take precedence. To improve affordability and uptake, strategies such as integrating other medical services into gHAT screening, offering modest community-level incentives, or providing village-level support when participation is high should be considered. Ultimately, integrating gHAT activities within broader health programs and surveillance systems will be essential to optimize resources and ensure long-term sustainability.

New treatments might offer potential solutions. With the recent introduction of fexinidazole, a 10-day oral treatment for both disease stages, only parasite confirmation is required. However WHO guidelines recommend supervised intake, therefore necessitating clinic services in practice.[32] Acoziborole, a single-dose oral treatment effective against both disease stages, received a positive scientific opinion from the EMA in February 2026 and could be a game changer by reducing costs and burden on both patients and the health care system [11]. It has the potential to enable treatment at the point of care and reduce the need for hospitalization. Further if ongoing screen and treat trials confirm its safety and feasibility, treatment could rely on serological testing alone, removing the need for parasitological confirmation the coming years.[11, 33-35]

To strengthen access, early diagnosis and surveillance, opportunities to decentralize gHAT screening should be investigated. For instance, it could be possible to integrate dried blood sample collection in primary care facilities and pharmacies where patients first seek care. Samples could be periodically picked up for centralized laboratory, with follow-up of positive cases and data coordination by the national control program.[36] Such an approach could leverage existing care-seeking patterns rather than requiring patients to travel to distant facilities.

## Conclusion

Achieving WHO’s 2030 goal of zero gHAT transmission will require more than free medication and diagnostics. Over 15% of rural minimum-wage households still face catastrophic health expenditures in passive screening and elimination strategies increasingly dependent on passive surveillance face fundamental challenges in rural, low-income settings. Increasingly, populations will encounter fewer cases, reducing disease awareness. Moreover, symptomatic individuals seek care first at nearby, low-cost facilities unable to diagnose gHAT, and few will travel more than one hour to expensive hospitals capable of diagnosis. Ensuring financial protection for the poorest and most remote populations will therefore be critical to sustain engagement in screening and treatment. Strengthening affordability, decentralizing diagnosis to facilities where patients already seek care, and integrating gHAT activities within broader health services will enhance access and efficiency. Continued investment in community awareness, transport support, and patient-centered approaches will be key to maintaining participation as disease prevalence declines. Ultimately, equitable and financially accessible care is essential for achieving and sustaining gHAT elimination.

## Supplementary information

Supporting Information 1 : Number of new reported gHAT cases during the last 10 years

Supporting Information 2 : Map of the study area

Supporting Information 3 : Table: Composition of FGDs and SSIs for each health zone

Supporting Information 4 : Table Knowledge on HAT characteristics by screening strategy

Supporting Information 5: Bar chart Awareness of HAT cases among participants by Screening Strategy

Supporting Information 6: Bar charts Reported symptoms prior to HAT by Screening Strategy

Supporting Information 7: Table Health Care Seeking Behavior Pathway of participants experiencing symptoms prior to screening

Supporting Information 8: Table Visits reported and travel time per type of health care structure

Supporting Information 9: Table Means of travel used for screening, confirmation and treatment by HAT screening outcome

Supporting Information 10: Table Travel time for screening, confirmation and treatment by HAT screening outcome

Supporting Information 10: Reported revenue loss due to screening, microscopy and treatment

Supporting Information 12: Summary of Statistical Tests

Supporting Information 13: GLM regression Analysis of the Probability of Incurring Out-of-Pocket Costs by visit

Supporting Information 14: GLM regression Analysis of the level of Out-of-Pocket Costs by visits reporting OOP

Supporting Information 15: Table Median OOP per cost type per type of health structure

Supporting Information 16: Table Median OOP per screening strategy per screening outcome

Supporting Information 17: Frequency of the structures visited by participants

Supporting Information 18: Interview guide – Individual semi-structured interviews

Supporting Information 19: Interview guide – Focus group discussions

Supporting Information 20: Survey « Evaluation des dépenses médicales et non-médicales THA »

## Acknowledgments

The authors acknowledge the staff that contributed to the data collection in the field during the quantitative and qualitative phases of the study: Komo Laurent, Belesi Blaise,Bakomeka Mpeya, Munsi Jean Fidele, Mubiala Modeste, Dr. Muley Serge, Musala Kiyula François, Kiyene Lvwanga Patience, Dr. Jean Claude Ndikisa, Malungu Tabita, Norbert Mikanza Mupindi, Muyaya Ngeyum Desty, Kalunga Mukitshi Dadou, Dr. Pacifique Nunda, Kimana Ndambu Junior, Mboma Kibolomvula Crispin, Arsène, Ruth Nzuzi. Further I would like to thank dr. Elena Nicco and dr. Catiane Vander Kelen for their valuable feedback when drafting the manuscript.

## Data availability

The data supporting this study are retained at the Institute of Tropical Medicine and are not openly accessible due to ethical privacy concerns. De-identified transcripts and survey results can be obtained from the research group upon request for further research purposes, under the condition that they will not be published in whole or in part. Data access requires approval through a motivated written request to the Institute of Tropical Medicine at.

## Footnotes

1 Primary care facilities (health posts, health centres, or reference centres) are typically staffed by nurses and generally do not have medical doctors on site.

2 Part A: Average cost chart: Inner ring = sum of component averages; Part B are coping strategies for OOP diag- nostic and treatment costs

3 CHE = catastrophic health expenditure, defined as OOP exceeding specified percentage of annual household income; MW = minimum wage household ($340/year); AW = average wage household ($1,900/year). Thresh- olds: MW 10% = $34; MW 25% = $85; AW 10% = $190; AW 25% = $474.

