## Supplementary information for "The Household Burden of Sleeping Sickness: Out-of-Pocket Costs for Diagnosis and Treatment"

### I. Supplementary information

**Supporting Information 1 : Number of new reported gHAT cases during the last 10 years**

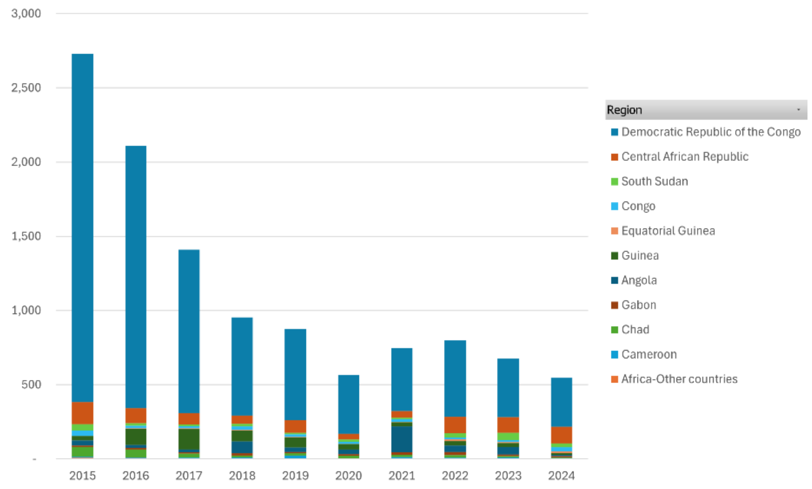

Reference: World Health Organization (WHO). Human African trypanosomiasis: Number of new reported cases of Human African trypanosomiasis (T.b. gambiense):2024 Geneva: WHO; 2024 [updated 01/07/2025; cited 2025 08/08/2025]. Available from: https://apps.who.int/neglected_diseases/ntddata/hat/hat.html

**Supporting Information 2 : Map of the study area**

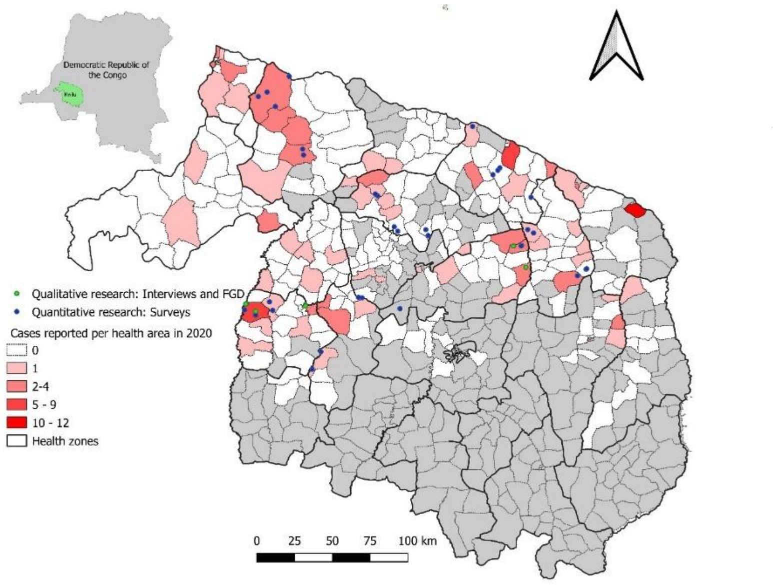

**Reference:** Snijders R. Study Area: Zenodo; 2024. doi: https://doi.org/10.5281/zenodo.12624925.

**Supporting Information 3 : Table: Composition of FGDs and SSIs for each health zone**

| **Participants FGD's and SSIs** | **HZ A** | **HZ B** |
|---|---|---|
| FGD Mixed: Health facility workers working in facilities without HAT screening | | 1 |
| FGD Mixed: Health facility workers working in facilities with HAT screening | | 1 |
| FGD Male, HAT endemic villages | 1 | 1 |
| FGD Female, HAT endemic villages | 1 | 1 |
| SSI Community Leader Male, HAT endemic villages | 1 | 1 |
| SSI PS Male and Female, HAT endemic villages | 6 | 1 |
| SSI AS Male and Female, HAT endemic village | 2 | 5 |
| **Total** | **11** | **11** |

**Supporting Information 4 : Table Knowledge on HAT characteristics by screening strategy**

| | Active (N=230) | Passive (N=214) | Overall (N=444) |
|---|---|---|---|
| **Knowledge on HAT** | | | |
| Mean (SD) | 2.87 (1.10) | 2.97 (1.23) | 2.91 (1.16) |
| Median [Min, Max] | 3.00 [0, 5.00] | 3.00 [0, 5.00] | 3.00 [0, 5.00] |
| **HAT characteristics reported** | | | |
| Sleep disorders | 148 (64.3%) | 135 (63.1%) | 283 (63.7%) |
| Headaches | 134 (58.3%) | 128 (59.8%) | 262 (59.0%) |
| Fever | 115 (50.0%) | 107 (50.0%) | 222 (50.0%) |
| Fatigue | 68 (29.6%) | 59 (27.6%) | 127 (28.6%) |
| Muscle and joint pain | 43 (18.7%) | 34 (15.9%) | 77 (17.3%) |
| Behavioral disorders | 32 (13.9%) | 39 (18.2%) | 71 (16.0%) |
| Other | 24 (10.4%) | 40 (18.7%) | 64 (14.4%) |
| Tse tse flies | 31 (13.5%) | 19 (8.9%) | 50 (11.3%) |
| Pruritis, itching | 26 (11.3%) | 14 (6.5%) | 40 (9.0%) |
| Loss of appetite and/or weight loss | 11 (4.8%) | 16 (7.5%) | 27 (6.1%) |
| Neurological disorders | 8 (3.5%) | 15 (7.0%) | 23 (5.2%) |
| Nothing | 9 (3.9%) | 8 (3.7%) | 17 (3.8%) |
| A fatal disease | 2 (0.9%) | 13 (6.1%) | 15 (3.4%) |
| Vomiting, dizziness, nausea, diarrhea | 4 (1.7%) | 7 (3.3%) | 11 (2.5%) |
| Low back pain, lumbar pain | 3 (1.3%) | 6 (2.8%) | 9 (2.0%) |
| Loss of consciousness, coma | 6 (2.6%) | 3 (1.4%) | 9 (2.0%) |
| Ganglions | 4 (1.7%) | 0 (0%) | 4 (0.9%) |

**Supporting Information 5: Bar chart Awareness of HAT cases among participants by Screening Strategy**

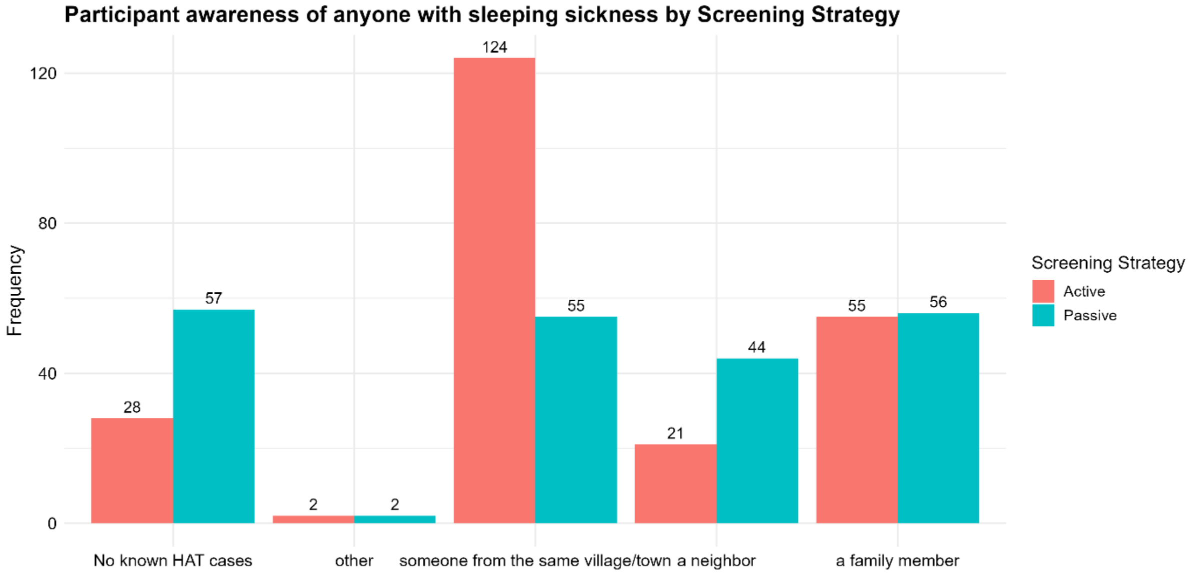

**Supporting Information 6: Bar charts Reported symptoms prior to HAT by Screening Strategy**

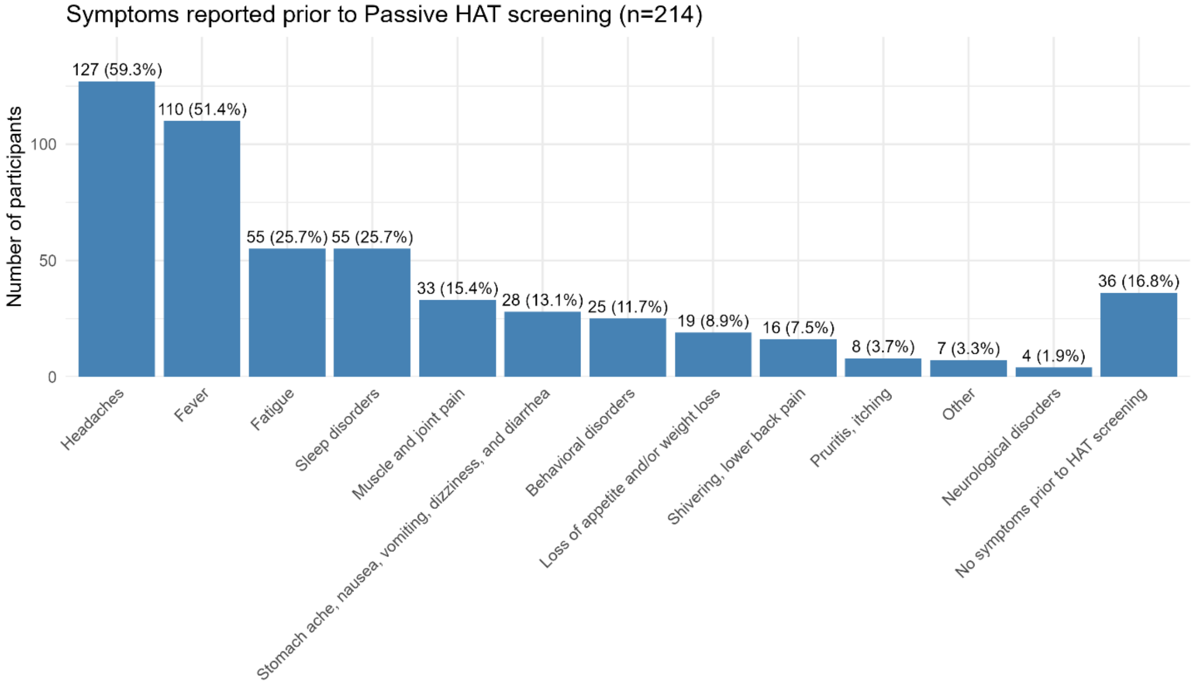

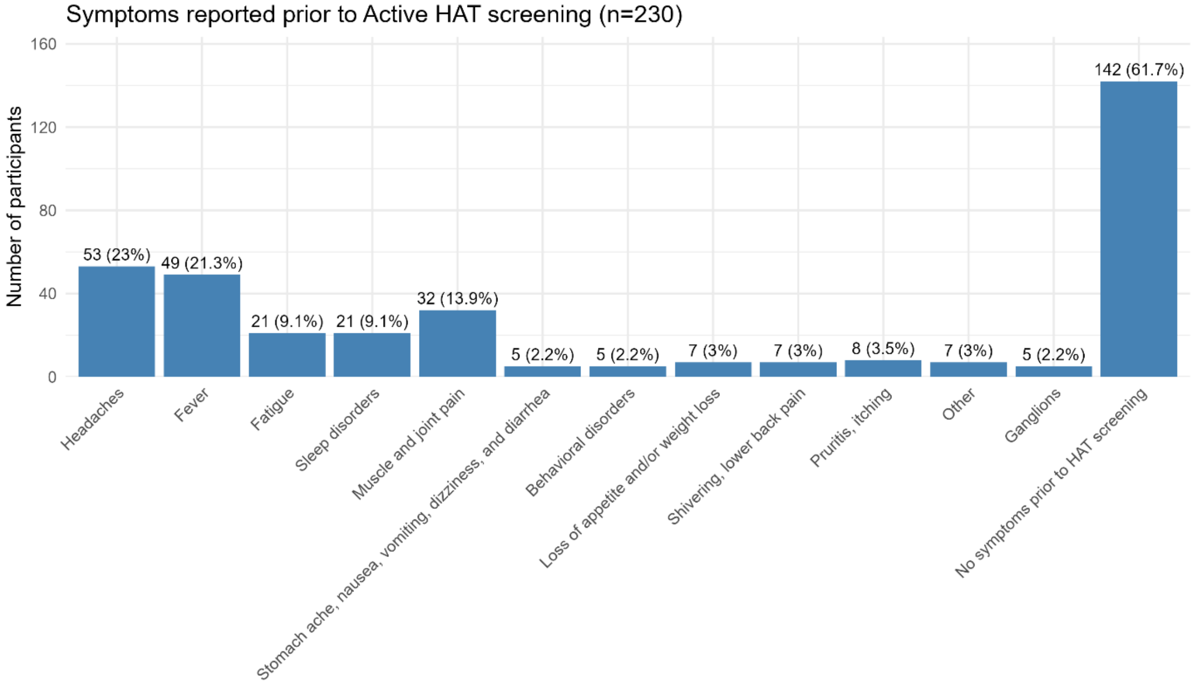

#### Supporting Information 7: Table Health Care Seeking Behavior Pathway of participants experiencing symptoms prior to screening

| Type of health structure | 1St | 2nd | 3rd | 4th | 5th | 6th | 7th | 8th | 9th |
|---|---|---|---|---|---|---|---|---|---|
| Traditional practitioner | 5(3%) | 57(33%) | 79(45%) | 19(11%) | 10(6%) | 2(1%) | 2(1%) | 0(0%) | 0(0%) |
| Pharmacy | 71(41%) | 4(2%) | 58(33%) | 38(22%) | 3(2%) | 0(0%) | 0(0%) | 0(0%) | 0(0%) |
| Primary care facility | 79(45%) | 41(24%) | 9(5%) | 41(24%) | 3(2%) | 1(1%) | 0(0%) | 0(0%) | 0(0%) |
| Hospital | 16(9%) | 58(33%) | 20(11%) | 62(36%) | 17(10%) | 1(1%) | 0(0%) | 0(0%) | 0(0%) |
| Private medical practices | 2(1%) | 3(2%) | 0(0%) | 1(1%) | 139(80%) | 27(16%) | 2(1%) | 0(0%) | 0(0%) |
| Mobile team with a vehicle | 1(1%) | 7(4%) | 4(2%) | 10(6%) | 1(1%) | 142(82%) | 9(5%) | 0(0%) | 0(0%) |
| Mobile team with motorcycles | 0(0%) | 2(1%) | 2(1%) | 1(1%) | 1(1%) | 1(1%) | 161(93%) | 6(3%) | 0(0%) |
| Screening, diagnosis, treatment, and control center (CDTC) | 0(0%) | 2(1%) | 1(1%) | 2(1%) | 0(0%) | 0(0%) | 0(0%) | 168(97%) | 1(1%) |
| Other | 0(0%) | 0(0%) | 1(1%) | 0(0%) | 0(0%) | 0(0%) | 0(0%) | 0(0%) | 173(99%) |

#### Supporting Information 8: Table Visits reported and travel time per type of health care structure

| Structure type | Traveltime | Frequency | % |
|---|---|---|---|
| Pharmacy | 0 | 12 | 12% |
| Pharmacy | 1hr or less | 88 | 87% |
| Pharmacy | half a day or less | 1 | 1% |
| **Pharmacy Total** | | **101** | **100%** |
| Health post - Health center - Referral center | 0 | 39 | 14% |
| Health post - Health center - Referral center | 1hr or less | 160 | 57% |
| Health post - Health center - Referral center | half a day or less | 73 | 26% |
| Health post - Health center - Referral center | 1 day | 7 | 3% |
| Health post - Health center - Referral center | 2 days | 1 | 0% |
| **Health post - Health center - Referral center Total** | | **280** | **100%** |
| Secondary hospital - General referral hospital | 0 | 4 | 2% |
| Secondary hospital - General referral hospital | 1hr or less | 78 | 48% |
| Secondary hospital - General referral hospital | half a day or less | 56 | 35% |
| Secondary hospital - General referral hospital | 1 day | 22 | 14% |
| Secondary hospital - General referral hospital | 2 days | 1 | 1% |
| **Secondary hospital - General referral hospital Total** | | **161** | **100%** |
| A mobile team with a vehicle | 0 | 88 | 47% |
| A mobile team with a vehicle | 1hr or less | 92 | 49% |
| A mobile team with a vehicle | half a day or less | 8 | 4% |
| A mobile team with a vehicle | 1 day | 1 | 1% |
| **A mobile team with a vehicle Total** | | **189** | **100%** |
| A mobile team with motorcycles | 0 | 15 | 37% |
| A mobile team with motorcycles | 1hr or less | 25 | 61% |
| A mobile team with motorcycles | half a day or less | 1 | 2% |
| **A mobile team with motorcycles Total** | | **41** | **100%** |
| Other | 0 | 2 | 100% |
| **Other Total** | | **2** | **100%** |
| Private center - Private medical practice | 0 | 1 | 50% |
| Private center - Private medical practice | half a day or less | 1 | 50% |
| **Private center - Private medical practice Total** | | **2** | **100%** |
| Traditional practitioners | 1hr or less | 5 | 45% |
| Traditional practitioners | 1 day | 5 | 45% |
| Traditional practitioners | 2 days | 1 | 9% |
| **Traditional practitioners Total** | | **11** | **100%** |
| Screening, diagnosis, treatment, and monitoring center (CDTC) | half a day or less | 5 | 38% |
| Screening, diagnosis, treatment, and monitoring center (CDTC) | 1hr or less | 6 | 46% |
| Screening, diagnosis, treatment, and monitoring center (CDTC) | 1 day | 1 | 8% |
| Screening, diagnosis, treatment, and monitoring center (CDTC) | 2 days | 1 | 8% |
| **Screening, diagnosis, treatment, and monitoring center (CDTC) Total** | | **13** | **100%** |
| **Grand Total** | | **800** | |

**Supporting Information 9: Table Means of travel used for screening, confirmation and treatment by HAT screening outcome**

| Means of travel | Serology negative | | | Serology result unknown | | | Serology positive with microscopy not done/unknown | | | Serology performed with microscopy negative | | | Confirmed HAT case | | | Overall | | |
|---|---|---|---|---|---|---|---|---|---|---|---|---|---|---|---|---|---|---|
| | AS | PS | Total | AS | PS | Total | AS | PS | Total | AS | PS | Total | AS | PS | Total | AS | PS | Total |
| **Screening** | **(N=128)** | **(N=134)** | **(N=262)** | **(N=10)** | **(N=11)** | **(N=21)** | **(N=13)** | **(N=9)** | **(N=22)** | **(N=61)** | **(N=32)** | **(N=93)** | **(N=18)** | **(N=28)** | **(N=46)** | **(N=230)** | **(N=214)** | **(N=444)** |
| Screened without travel | 103 (80.5%) | 13 (9.7%) | 116 (44.3%) | 10 (100%) | 1 (9.1%) | 11 (52.4%) | 13 (100%) | 3 (33.3%) | 16 (72.7%) | 46 (75.4%) | 15 (46.9%) | 61 (65.6%) | 17 (94.4%) | 10 (35.7%) | 27 (58.7%) | 189 (82.2%) | 42 (19.6%) | 231 (52.0%) |
| On foot | 19 (14.8%) | 81 (60.4%) | 100 (38.2%) | 0 (0%) | 6 (54.5%) | 6 (28.6%) | 0 (0%) | 5 (55.6%) | 5 (22.7%) | 15 (24.6%) | 12 (37.5%) | 27 (29.0%) | 1 (5.6%) | 16 (57.1%) | 17 (37.0%) | 35 (15.2%) | 120 (56.1%) | 155 (34.9%) |
| By bike | 1 (0.8%) | 8 (6.0%) | 9 (3.4%) | 0 (0%) | 0 (0%) | 0 (0%) | 0 (0%) | 1 (11.1%) | 1 (4.5%) | 0 (0%) | 1 (3.1%) | 1 (1.1%) | 0 (0%) | 0 (0%) | 0 (0%) | 1 (0.4%) | 10 (4.7%) | 11 (2.5%) |
| By motorcycle | 3 (2.3%) | 31 (23.1%) | 34 (13.0%) | 0 (0%) | 4 (36.4%) | 4 (19.0%) | 0 (0%) | 0 (0%) | 0 (0%) | 0 (0%) | 4 (12.5%) | 4 (4.3%) | 0 (0%) | 2 (7.1%) | 2 (4.3%) | 3 (1.3%) | 41 (19.2%) | 44 (9.9%) |
| Other[1] | 2 (1.6%) | 1 (0.7%) | 3 (1.1%) | 0 (0%) | 0 (0%) | 0 (0%) | 0 (0%) | 0 (0%) | 0 (0%) | 0 (0%) | 0 (0%) | 0 (0%) | 0 (0%) | 0 (0%) | 0 (0%) | 2 (0.9%) | 1 (0.5%) | 3 (0.7%) |
| **Confirmation** | | | | | | | | | | **(N=61)** | **(N=32)** | **(N=93)** | **(N=18)** | **(N=28)** | **(N=46)** | **(N=79)** | **(N=60)** | **(N=139)** |
| On the spot follow-up testing | | | | | | | | | | 54 (88.5%) | 26 (81.3%) | 80 (86.0%) | 15 (83.3%) | 26 (92.9%) | 41 (89.1%) | 69 (87.3%) | 52 (86.7%) | 121 (87.0%) |
| Follow-up testing later at home | | | | | | | | | | 1 (1.6%) | 6 (18.8%) | 7 (7.5%) | 1 (5.6%) | 0 (0%) | 1 (2.2%) | 2 (2.5%) | 6 (10,0%) | 8 (5.7%) |
| On foot | | | | | | | | | | 5 (8.2%) | 0 (0%) | 5 (5.4%) | 1 (5.6%) | 2 (7.1%) | 3 (6.5%) | 6 (7.6%) | 2 (3,3%) | 8 (5.7%) |
| By motorcycle | | | | | | | | | | 1 (1.6%) | 0 (0%) | 1 (1.1%) | 1 (5.6%) | 0 (0%) | 1 (2.2%) | 2 (2.5%) | 0 (0%) | 2 (1.6%) |
| **Treatment** | | | | | | | | | | | | | **(N=18)** | **(N=28)** | **(N=46)** | **(N=18)** | **(N=28)** | **(N=46)** |
| Treated without travel | | | | | | | | | | | | | 4 (22.2%) | 16 (57.1%) | 20 (43.5%) | 4 (22.2%) | 16 (57.1%) | 20 (43.5%) |
| On foot | | | | | | | | | | | | | 9 (50.0%) | 8 (28.6%) | 17 (37.0%) | 9 (50.0%) | 8 (28.6%) | 17 (37.0%) |
| By motorcycle | | | | | | | | | | | | | 4 (22.2%) | 3 (10.7%) | 7 (15.2%) | 4 (22.2%) | 3 (10.7%) | 7 (15.2%) |
| By car | | | | | | | | | | | | | 1 (5.6%) | 0 (0%) | 1 (2.2%) | 1 (5.6%) | 0 (0%) | 1 (2.2%) |
| Other | | | | | | | | | | | | | 0 (0%) | 1 (3.6%) | 1 (2.2%) | 0 (0%) | 1 (3.6%) | 1 (2.2%) |

[1] Motorcycle of the health district or the HAT screening mobile team

**Supporting Information 10: Table Travel time for screening, confirmation and treatment by HAT screening outcome**

| Travel time | Serology negative | | | Serology result unknown | | | Serology positive with microscopy not done/unknown | | | Serology performed with microscopy negative | | | Confirmed HAT case | | | Overall | | |
|---|---|---|---|---|---|---|---|---|---|---|---|---|---|---|---|---|---|---|
| | AS | PS | Total | AS | PS | Total | AS | PS | Total | AS | PS | Total | AS | PS | Total | AS | PS | Total |
| **Screening** | **(N=128)** | **(N=134)** | **(N=262)** | **(N=10)** | **(N=11)** | **(N=21)** | **(N=10)** | **(N=11)** | **(N=21)** | **(N=61)** | **(N=32)** | **(N=93)** | **(N=18)** | **(N=28)** | **(N=46)** | **(N=230)** | **(N=214)** | **(N=444)** |
| No travel time | 103 (80.5%) | 13 (9.7%) | 116 (44.3%) | 10 (100%) | 1 (9.1%) | 11 (52.4%) | 13 (100%) | 3 (33.3%) | 16 (72.7%) | 46 (75.4%) | 15 (46.9%) | 61 (65.6%) | 17 (94.4%) | 10 (35.7%) | 27 (58.7%) | 189 (82.2%) | 42 (19.6%) | 231 (52.0%) |
| Less than 30 minutes | 14 (10.9%) | 63 (47.0%) | 77 (29.4%) | 0 (0%) | 5 (45.5%) | 5 (23.8%) | 0 (0%) | 4 (44.4%) | 4 (18.2%) | 8 (13.1%) | 9 (28.1%) | 17 (18.3%) | 1 (5.6%) | 4 (14.3%) | 5 (10.9%) | 23 (10.0%) | 85 (39.7%) | 108 (24.3%) |
| Less than an hour | 2 (1.6%) | 31 (23.1%) | 33 (12.6%) | 0 (0%) | 4 (36.4%) | 4 (19.0%) | 0 (0%) | 1 (11.1%) | 1 (4.5%) | 5 (8.2%) | 5 (15.6%) | 10 (10.8%) | 0 (0%) | 4 (14.3%) | 4 (8.7%) | 7 (3.0%) | 45 (21.0%) | 52 (11.7%) |
| Less than 2 hours | 4 (3.1%) | 15 (11.2%) | 19 (7.3%) | 0 (0%) | 1 (9.1%) | 1 (4.8%) | 0 (0%) | 0 (0%) | 0 (0%) | 2 (3.3%) | 0 (0%) | 2 (2.2%) | 0 (0%) | 3 (10.7%) | 3 (6.5%) | 6 (2.6%) | 19 (8.9%) | 25 (5.6%) |
| Less than 4 hours | 2 (1.6%) | 8 (6.0%) | 10 (3.8%) | 0 (0%) | 0 (0%) | 0 (0%) | 0 (0%) | 0 (0%) | 0 (0%) | 0 (0%) | 3 (9.4%) | 3 (3.2%) | 0 (0%) | 5 (17.9%) | 5 (10.9%) | 2 (0.9%) | 16 (7.5%) | 18 (4.1%) |
| Less than 8 hours | 2 (1.6%) | 3 (2.2%) | 5 (1.9%) | 0 (0%) | 0 (0%) | 0 (0%) | 0 (0%) | 1 (11.1%) | 1 (4.5%) | 0 (0%) | 0 (0%) | 0 (0%) | 0 (0%) | 1 (3.6%) | 1 (2.2%) | 2 (0.9%) | 5 (2.3%) | 7 (1.6%) |
| Less than a day | 1 (0.8%) | 1 (0.7%) | 2 (0.8%) | 0 (0%) | 0 (0%) | 0 (0%) | 0 (0%) | 0 (0%) | 0 (0%) | 0 (0%) | 0 (0%) | 0 (0%) | 0 (0%) | 0 (0%) | 0 (0%) | 1 (0.4%) | 1 (0.5%) | 2 (0.5%) |
| More than one day, I had to spend the night somewhere along the way. | 0 (0%) | 0 (0%) | 0 (0%) | 0 (0%) | 0 (0%) | 0 (0%) | 0 (0%) | 0 (0%) | 0 (0%) | 0 (0%) | 0 (0%) | 0 (0%) | 0 (0%) | 1 (3.6%) | 1 (2.2%) | 0 (0%) | 1 (0.5%) | 1 (0.2%) |
| **Confirmation** | | | | | | | | | | **(N=61)** | **(N=32)** | **(N=93)** | **(N=18)** | **(N=28)** | **(N=46)** | **(N=79)** | **(N=60)** | **(N=139)** |
| No additional travel required | | | | | | | | | | 55 (90.2%) | 32 (100%) | 87 (93.5%) | 16 (88.9%) | 26 (92.9%) | 42 (91.3%) | 71 (89.9%) | 58 (96.7%) | 129 (92.8%) |
| Less than 30 minutes | | | | | | | | | | 4 (6.6%) | 0 (0%) | 4 (4.3%) | 0 (0%) | 0 (0%) | 0 (0%) | 4 (5.1%) | | 4 (2.9%) |
| Less than an hour | | | | | | | | | | 1 (1.6%) | 0 (0%) | 1 (1.1%) | 0 (0%) | 0 (0%) | 0 (0%) | 1 (1.2%) | | 1 (0.7%) |
| Less than 2 hours | | | | | | | | | | 1 (1.6%) | 0 (0%) | 1 (1.1%) | 0 (0%) | 0 (0%) | 0 (0%) | 1 (1.2%) | | 1 (0.7%) |
| Less than 4 hours | | | | | | | | | | 0 (0%) | 0 (0%) | 0 (0%) | 1 (5.6%) | 1 (3.6%) | 2 (4.3%) | 1 (1.2%) | 1 (1.6%) | 2 (1.4%) |
| Less than 8 hours | | | | | | | | | | 0 (0%) | 0 (0%) | 0 (0%) | 1 (5.6%) | 1 (3.6%) | 2 (4.3%) | 1 (1.2%) | 1 (1.6%) | 2 (1.4%) |
| **Treatment** | | | | | | | | | | | | | **(N=18)** | **(N=28)** | **(N=46)** | **(N=18)** | **(N=28)** | **(N=46)** |
| No travel time | | | | | | | | | | | | | 4 (22.2%) | 16 (57.1%) | 20 (43.5%) | 4 (22.2%) | 16 (57.1%) | 20 (43.5%) |
| Less than 30 minutes | | | | | | | | | | | | | 0 (0%) | 3 (10.7%) | 3 (6.5%) | 0 (0%) | 3 (10.7%) | 3 (6.5%) |
| Less than an hour | | | | | | | | | | | | | 3 (16.7%) | 1 (3.6%) | 4 (8.7%) | 3 (16.7%) | 1 (3.6%) | 4 (8.7%) |
| Less than 2 hours | | | | | | | | | | | | | 2 (11.1%) | 1 (3.6%) | 3 (6.5%) | 2 (11.1%) | 1 (3.6%) | 3 (6.5%) |
| Less than 4 hours | | | | | | | | | | | | | 4 (22.2%) | 3 (10.7%) | 7 (15.2%) | 4 (22.2%) | 3 (10.7%) | 7 (15.2%) |
| Less than 8 hours | | | | | | | | | | | | | 3 (16.7%) | 2 (7.1%) | 5 (10.9%) | 3 (16.7%) | 2 (7.1%) | 5 (10.9%) |
| Less than a day | | | | | | | | | | | | | 2 (11.1%) | 1 (3.6%) | 3 (6.5%) | 2 (11.1%) | 1 (3.6%) | 3 (6.5%) |
| More than one day, I had to spend the night somewhere along the way. | | | | | | | | | | | | | 0 (0%) | 1 (3.6%) | 1 (2.2%) | 0 (0%) | 1 (3.6%) | 1 (2.2%) |

**Supporting Information 11: Reported revenue loss due to screening, microscopy and treatment**

| Revenu loss due to screening | Participants | % by Total |
|---|---|---|
| **No** | **203** | **46%** |
| Active | 148 | 33% |
| Passive | 55 | 12% |
| **Yes** | **241** | **54%** |
| Active | 82 | 18% |
| Passive | 159 | 36% |
| **Grand Total** | **444** | **100%** |
| **Revenu loss due to microscopy confirmation** | **Participants** | **% by Total** |
| **No** | **93** | **64%** |
| Active | 68 | 47% |
| Passive | 25 | 17% |
| **Yes** | **53** | **36%** |
| Active | 18 | 12% |
| Passive | 35 | 24% |
| **Grand Total** | **146** | **100%** |
| **Revenu loss due to treatment** | **Participants** | **% by Total** |
| **No** | **5** | **11%** |
| Active | 3 | 7% |
| Passive | 2 | 4% |
| **Yes** | **41** | **89%** |
| Active | 15 | 33% |
| Passive | 26 | 57% |
| **Grand Total** | **46** | **100%** |

**Supporting Information 12: Summary of Statistical Tests**

1. **Comparison Positive and Zero-OOP Proportions Between Screening strategies for visits and participants, including food costs**

| | | **Visits (n= 800)** | | | | | | **Participants (n=444)** | | | | | |
|---|---|---|---|---|---|---|---|---|---|---|---|---|---|
| **Strategy** | **Comparison** | **n OOP=0** | **n OOP >0** | **test** | **Chi-square** | **df** | **p_value** | **n OOP=0** | **n OOP >0** | **test** | **Chi-square** | **df** | **p_value** |
| AS | Zero OOP vs Positive OOP | 215 | 15 | Chi-square | 467.58 | 1 | p < 0.0001 | 126 | 104 | Chi-square | 83.25 | 1 | p < 0.0001 |
| PS | Zero OOP vs Positive OOP | 70 | 500 | | | | | 28 | 186 | Chi-square | | | |
| | | **285** | **515** | | | | | 154 | 290 | | | | |

2. **Comparison Positive and Zero-OOP Proportions Between Screening strategies for visits and participants, excluding food costs**

| | | **Visits (n= 800)** | | | | | | **Participants (n=444)** | | | | | |
|---|---|---|---|---|---|---|---|---|---|---|---|---|---|
| **Strategy** | **Comparison** | **n OOP=0** | **n OOP >0** | **test** | **Chi-square** | **df** | **p_value** | **n OOP=0** | **n OOP >0** | **test** | **Chi-square** | **df** | **p_value** |
| AS | Zero OOP vs Positive OOP | 223 | 7 | Chi-square | 449.89 | 1 | p < 0.0001 | 134 | 96 | Chi-square | 76.86 | 1 | p < 0.0001 |
| PS | Zero OOP vs Positive OOP | 90 | 480 | | | | | 37 | 177 | Chi-square | | | |
| | | **313** | **487** | | | | | 171 | 273 | | | | |

3. **Comparison of Median OOP Costs Between Screening Strategies Among Visits With Positive OOP, including food costs**

| | **Visits (n= 800)** | | | | | **Participants (n=444)** | | | | |
|---|---|---|---|---|---|---|---|---|---|---|
| **Comparison** | **AS** | **PS** | **test** | **MW** | **p_value** | **AS** | **PS** | **test** | **MW** | **p_value** |
| Median OOP AS & PS including food costs | 0.76 | 9.08 | Mann-Whitney U-test | 966 | p < 0.0001 | 12.14 | 20.51 | Mann-Whitney U-test | 8324.5 | 0.05 |
| Median OOP AS & PS excluding food costs | 1.01 | 7.57 | Mann-Whitney U-test | 342.5 | P<0.001 | 10.80 | 17.66 | Mann-Whitney U-test | 7509 | 0.11 |

4. **Comparison among confirmed cases of patients that were identified via AS vs PS, including food costs**

| | | Participants (n= 46) | | | | | |
|---|---|---|---|---|---|---|---|
| **Strategy** | **Comparison** | **n OOP=0** | **n OOP >0** | **test** | **p_value** | **95% CI** | **Odds ratio** |
| AS | Zero OOP vs Positive OOP | 4 | 14 | Fisher's exact test | p = 0.019 | (1.13-Inf) | inf |
| PS | Zero OOP vs Positive OOP | 0 | 28 | | | | |
| | | **4** | **42** | | | | |

**5. Comparison among confirmed cases of patients that were identified via AS vs PS, excluding food costs**

| | | Participants (n= 46) | | | | | |
|---|---|---|---|---|---|---|---|
| **Strategy** | **Comparison** | **n OOP=0** | **n OOP >0** | **test** | **p_value** | **95% CI** | **Odds ratio** |
| AS | Zero OOP vs Positive OOP | 7 | 11 | Fisher's exact test | P=0.018 | (1.239 - 89.391) | 7.857 |
| PS | Zero OOP vs Positive OOP | 2 | 26 | | | | |
| | | **9** | **37** | | | | |

**6. Comparison of Median OOP Costs Between Screening Strategies Among Confirmed Cases with Positive OOP**

| | Participants (n=46) | | | | |
|---|---|---|---|---|---|
| **Comparison** | **AS (median)** | **PS (median)** | **test** | **MW** | **p_value** |
| Median OOP AS & PS including food costs | 9.84 | 24.52 | Mann-Whitney U-test | 92 | 0.006 |
| Median OOP AS & PS excluding food costs | 5.05 | 22.83 | Mann-Whitney U-test | 57 | 0.004 |

**Supporting Information 13: GLM regression Analysis of the Probability of Incurring Out-of-Pocket Costs by visit**
**Model:** Binomial logistic regression (N =800)
**Outcome variable**: No OOP versus OOP
**Predictor variables:** Type of health structure, Travel time, mode of transport, Screening outcome

Table1: Results Likelihood Ratio Tests for single-term deletions

| Predictor | df | Deviance | AIC | LRT | p-value |
|---|---|---|---|---|---|
| **Full model** | — | 449.54 | 473.54 | — | — |
| Type of health structure | 4 | 848.51 | 864.51 | 398.97 | <0.00001 |
| Travel time | 2 | 455.26 | 475.26 | 5.73 | 0.057 |
| Mode of transport | 1 | 450.23 | 472.23 | 0.69 | 0.405 |
| Screening outcome | 4 | 454.43 | 470.43 | 4.90 | 0.298 |

Table2: Adjusted odds ratios (OR) and 95% confidence intervals (CI) for predictors of out-of-pocket (excluding screening outcome and mode of transport)

| Predictor | OR | 95% CI | p-value | Comment |
|---|---|---|---|---|
| **Health structure type (ref: Mobile team: Vehicle or motorcycle)** | | | | |
| Pharmacy | 442.23 | 124.33-1572.63 | <0.001 | Strongest predictor |
| Primary care facility | 42.63 | 23.05-78.77 | <0.001 | Large effect |
| Hospital | 241.82 | 84.73-690.03 | <0.001 | Very strong effect |
| Other types | 40.45 | 12.62-129.14 | <0.001 | Large effect |
| **Travel time (ref: No travel reported)** | | | | |
| 1 hour or less | 4.17 | 2.27-7.64 | <0.001 | Moderate increase in odds |
| More than 1 hour | 9.86 | 4.42-22.00 | <0.001 | Moderate increase in odds |

**Supporting Information 14: GLM regression Analysis of the level of Out-of-Pocket Costs by visits reporting OOP**
**Model:** Gaussian GLM with log link (N = 515)
**Outcome variable**: Total OOP costs including food costs (USD) among participants with OOP > 0
**Predictor variables:** Type of health structure, Travel time, mode of transport, Screening outcome

Table1: Results Likelihood Ratio Tests for single-term deletions

| Predictor | df | Deviance | AIC | Scaled dev. | p-value |
|---|---|---|---|---|---|
| **Full model** | — | 381,732 | 4891 | — | — |
| Type of health structure | 4 | 501,162 | 5023 | 140.19 | <0.001 |
| Travel time | 2 | 384.923 | 4891 | 4.29 | 0.117 |
| Mode of transport | 1 | 382.617 | 4890 | 1.19 | 0.275 |
| Screening outcome | 4 | 386.728 | 4890 | 6.70 | 0.153 |

Table2: Exponentiated coefficients (cost multipliers) and 95% confidence intervals for predictors of OOP cost level

**Model:** Gaussian GLM with log link (N = 515)
**Outcome variable**: Total OOP costs (USD) among participants with OOP > 0
**Predictor variables:** Type of health structure

| Predictor | Exp(β) | 95% CI | p-value | Interpretation |
|---|---|---|---|---|
| **Health structure type (ref: Mobile team)[2]** | | | | |
| Pharmacy | 2.19 | 0.00 – 2314 | 0.825 | Not significant |
| Primary care facility | 4.91 | 0.01 – 4697 | 0.650 | Not significant |
| Hospital | 21.12 | 0.02 – 19855 | 0.383 | Not significant |
| Other types | 11.92 | 0.01 – 11811 | 0.480 | Not significant |

[2] Note: Type of health structure was a highly significant predictor of cost level ($p < 0.001$). Individual coefficient p-values are not significant due to high variance in OOP costs and small sample sizes within mobile team reference category. Exp(β) represents the multiplicative effect on costs relative to mobile team screening.

**Supporting Information 15: Table Median OOP per cost type per type of health structure**

| StructTyp | variable | n_total | n_nonzero | Min_Value | Q1 | Median | Q3 | Max_Value | IQR |
|---|---|---|---|---|---|---|---|---|---|
| A mobile team with a vehicle | Medical OOP | 189 | 4.00 | 0.50 | 0.88 | 1.26 | 2.40 | 5.05 | 1.51 |
| A mobile team with a vehicle | Medical OOP: Consumables | 2 | 2.00 | 1.01 | 2.02 | 3.03 | 4.04 | 5.05 | 2.02 |
| A mobile team with a vehicle | Medical OOP: Laboratory | 1 | 1.00 | 0.50 | 0.50 | 0.50 | 0.50 | 0.50 | - |
| A mobile team with a vehicle | Medical OOP: Medication | 1 | 1.00 | 1.51 | 1.51 | 1.51 | 1.51 | 1.51 | - |
| A mobile team with a vehicle | Non-Medical OOP | 189 | 10.00 | 0.10 | 0.25 | 0.25 | 0.95 | 10.09 | 0.69 |
| A mobile team with a vehicle | Non-Medical OOP: Food | 8 | 8.00 | 0.10 | 0.25 | 0.25 | 2.27 | 10.09 | 2.02 |
| A mobile team with a vehicle | Non-Medical OOP: Other | 2 | 2.00 | 0.25 | 0.44 | 0.63 | 0.82 | 1.01 | 0.38 |
| A mobile team with a vehicle | Non-Medical OOP: Transport | 1 | 1.00 | 0.20 | 0.20 | 0.20 | 0.20 | 0.20 | - |
| A mobile team with a vehicle | Total OOP | 189 | 14.00 | 0.10 | 0.25 | 0.63 | 1.39 | 10.09 | 1.14 |
| A mobile team with a vehicle | Total OOP excluding food related costs | 189 | 7.00 | 0.20 | 0.38 | 1.01 | 1.26 | 5.05 | 0.88 |
| A mobile team with motorcycles | Non-Medical OOP | 41 | 1.00 | 2.02 | 2.02 | 2.02 | 2.02 | 2.02 | - |
| A mobile team with motorcycles | Non-Medical OOP: Food | 1 | 1.00 | 2.02 | 2.02 | 2.02 | 2.02 | 2.02 | - |
| A mobile team with motorcycles | Total OOP | 41 | 1.00 | 2.02 | 2.02 | 2.02 | 2.02 | 2.02 | - |
| A mobile team with motorcycles | Total OOP excluding food related costs | 41 | - | - | - | - | - | - | - |
| Health post - Health center - Referral center | Medical OOP | 280 | 193.00 | 0.25 | 3.03 | 5.15 | 8.58 | 76.08 | 5.55 |
| Health post - Health center - Referral center | Medical OOP: Consumables | 183 | 183.00 | 0.50 | 2.02 | 3.53 | 5.05 | 32.80 | 3.03 |
| Health post - Health center - Referral center | Medical OOP: Hospitalization | 17 | 17.00 | 0.50 | 2.52 | 3.53 | 7.57 | 10.60 | 5.05 |
| Health post - Health center - Referral center | Medical OOP: Laboratory | 55 | 55.00 | 0.10 | 0.50 | 1.01 | 1.64 | 22.70 | 1.14 |
| Health post - Health center - Referral center | Medical OOP: Medication | 78 | 78.00 | 0.25 | 2.52 | 3.53 | 6.05 | 22.70 | 3.53 |
| Health post - Health center - Referral center | Medical OOP: Other | 10 | 10.00 | 0.10 | 0.25 | 0.30 | 0.50 | 50.45 | 0.25 |
| Health post - Health center - Referral center | Medical OOP: Other purchases | 5 | 5.00 | 0.76 | 0.76 | 1.01 | 12.61 | 50.45 | 11.86 |
| Health post - Health center - Referral center | Non-Medical OOP | 280 | 171.00 | 0.10 | 0.63 | 1.51 | 4.04 | 58.02 | 3.41 |
| Health post - Health center - Referral center | Non-Medical OOP: Accommodation | 2 | 2.00 | 1.26 | 2.71 | 4.16 | 5.61 | 7.06 | 2.90 |
| Health post - Health center - Referral center | Non-Medical OOP: Food | 140 | 140.00 | 0.10 | 0.73 | 1.51 | 3.09 | 50.45 | 2.36 |
| Health post - Health center - Referral center | Non-Medical OOP: Other | 22 | 22.00 | 0.25 | 0.25 | 0.63 | 2.31 | 15.14 | 2.06 |
| Health post - Health center - Referral center | Non-Medical OOP: Transport | 59 | 59.00 | 0.25 | 0.50 | 1.01 | 3.28 | 22.70 | 2.77 |
| Health post - Health center - Referral center | Total OOP | 280 | 223.00 | 0.25 | 3.36 | 6.56 | 11.00 | 128.05 | 7.64 |
| Health post - Health center - Referral center | Total OOP excluding food related costs | 280 | 210.00 | 0.25 | 3.03 | 5.32 | 9.04 | 77.60 | 6.02 |
| Other | Medical OOP | 2 | 1.00 | 2.52 | 2.52 | 2.52 | 2.52 | 2.52 | - |
| Other | Medical OOP: Consumables | 1 | 1.00 | 2.52 | 2.52 | 2.52 | 2.52 | 2.52 | - |
| Other | Total OOP | 2 | 1.00 | 2.52 | 2.52 | 2.52 | 2.52 | 2.52 | - |
| Other | Total OOP excluding food related costs | 2 | 1.00 | 2.52 | 2.52 | 2.52 | 2.52 | 2.52 | - |
| Other | Total OOP excluding food related costs | 2 | - | - | 0.63 | 1.26 | 1.89 | 3.78 | 2.52 |
| Pharmacy | Medical OOP | 101 | 97.00 | 0.25 | 1.01 | 2.52 | 5.05 | 25.23 | 4.04 |
| Pharmacy | Medical OOP: Consumables | 9 | 9.00 | 2.52 | 3.53 | 7.57 | 10.09 | 25.23 | 6.56 |
| Pharmacy | Medical OOP: Medication | 88 | 88.00 | 0.25 | 0.76 | 2.14 | 5.05 | 25.23 | 4.29 |
| Pharmacy | Non-Medical OOP | 101 | 3.00 | 3.53 | 4.29 | 5.05 | 6.31 | 7.57 | 2.02 |
| Pharmacy | Non-Medical OOP: Accommodation | 1 | 1.00 | 1.01 | 1.01 | 1.01 | 1.01 | 1.01 | - |
| Pharmacy | Non-Medical OOP: Food | 3 | 3.00 | 0.50 | 2.27 | 4.04 | 4.54 | 5.05 | 2.27 |
| Pharmacy | Non-Medical OOP: Transport | 2 | 2.00 | 2.52 | 2.65 | 2.77 | 2.90 | 3.03 | 0.25 |
| Pharmacy | Total OOP | 101 | 98.00 | 0.25 | 1.01 | 2.52 | 5.05 | 25.23 | 4.04 |

| | | | | | | | | | |
|---|---|---|---|---|---|---|---|---|---|
| Pharmacy | Total OOP excluding food related costs | 101 | 98.00 | 0.25 | 1.01 | 2.52 | 5.05 | 25.23 | 4.04 |
| Private center - Private medical practice | Medical OOP | 2 | 2.00 | 14.13 | 15.01 | 15.89 | 16.78 | 17.66 | 1.77 |
| Private center - Private medical practice | Medical OOP: Consumables | 2 | 2.00 | 5.05 | 6.31 | 7.57 | 8.83 | 10.09 | 2.52 |
| Private center - Private medical practice | Medical OOP: Laboratory | 1 | 1.00 | 2.52 | 2.52 | 2.52 | 2.52 | 2.52 | - |
| Private center - Private medical practice | Medical OOP: Medication | 2 | 2.00 | 6.56 | 6.81 | 7.06 | 7.32 | 7.57 | 0.50 |
| Private center - Private medical practice | Non-Medical OOP | 2 | 1.00 | 7.57 | 7.57 | 7.57 | 7.57 | 7.57 | - |
| Private center - Private medical practice | Non-Medical OOP: Food | 1 | 1.00 | 7.57 | 7.57 | 7.57 | 7.57 | 7.57 | - |
| Private center - Private medical practice | Total OOP | 2 | 2.00 | 17.66 | 18.67 | 19.68 | 20.69 | 21.70 | 2.02 |
| Private center - Private medical practice | Total OOP excluding food related costs | 2 | 2.00 | 14.13 | 15.01 | 15.89 | 16.78 | 17.66 | 1.77 |
| Private center - Private medical practice | Total OOP excluding food related costs | 2 | 14.13 | 12.36 | 15.01 | 15.89 | 16.78 | 19.42 | 17.66 |
| Screening, diagnosis, treatment, and monitoring center (CDTC) | Medical OOP | 13 | 6.00 | 6.05 | 6.12 | 6.94 | 8.32 | 10.09 | 2.21 |
| Screening, diagnosis, treatment, and monitoring center (CDTC) | Medical OOP: Consumables | 1 | 1.00 | 8.07 | 8.07 | 8.07 | 8.07 | 8.07 | - |
| Screening, diagnosis, treatment, and monitoring center (CDTC) | Medical OOP: Hospitalization | 4 | 4.00 | 6.05 | 6.24 | 6.94 | 8.20 | 10.09 | 1.96 |
| Screening, diagnosis, treatment, and monitoring center (CDTC) | Medical OOP: Laboratory | 1 | 1.00 | 0.50 | 0.50 | 0.50 | 0.50 | 0.50 | - |
| Screening, diagnosis, treatment, and monitoring center (CDTC) | Medical OOP: Medication | 1 | 1.00 | 6.05 | 6.05 | 6.05 | 6.05 | 6.05 | - |
| Screening, diagnosis, treatment, and monitoring center (CDTC) | Non-Medical OOP | 13 | 7.00 | 1.26 | 6.31 | 9.33 | 38.09 | 90.82 | 31.79 |
| Screening, diagnosis, treatment, and monitoring center (CDTC) | Non-Medical OOP: Food | 4 | 4.00 | 4.29 | 8.64 | 23.97 | 43.52 | 60.54 | 34.88 |
| Screening, diagnosis, treatment, and monitoring center (CDTC) | Non-Medical OOP: Other | 2 | 2.00 | 0.50 | 7.95 | 15.39 | 22.83 | 30.27 | 14.88 |
| Screening, diagnosis, treatment, and monitoring center (CDTC) | Non-Medical OOP: Transport | 5 | 5.00 | 1.26 | 5.05 | 6.05 | 6.56 | 27.75 | 1.51 |
| Screening, diagnosis, treatment, and monitoring center (CDTC) | Total OOP | 13 | 9.00 | 1.26 | 10.09 | 13.62 | 16.65 | 90.82 | 6.56 |
| Screening, diagnosis, treatment, and monitoring center (CDTC) | Total OOP excluding food related costs | 13 | 9.00 | 1.26 | 8.58 | 11.10 | 13.62 | 30.27 | 5.05 |
| Secondary hospital - General referral hospital | Medical OOP | 161 | 147.00 | 0.50 | 11.83 | 22.20 | 36.07 | 376.14 | 24.24 |
| Secondary hospital - General referral hospital | Medical OOP: Consumables | 141 | 141.00 | 0.76 | 2.27 | 6.05 | 13.62 | 85.77 | 11.35 |
| Secondary hospital - General referral hospital | Medical OOP: Hospitalization | 39 | 39.00 | 1.26 | 3.78 | 6.05 | 10.09 | 30.27 | 6.31 |
| Secondary hospital - General referral hospital | Medical OOP: Laboratory | 74 | 73.00 | 0.25 | 1.01 | 1.77 | 3.53 | 28.25 | 2.52 |
| Secondary hospital - General referral hospital | Medical OOP: Medication | 108 | 108.00 | 0.25 | 7.57 | 12.61 | 20.81 | 100.91 | 13.24 |
| Secondary hospital - General referral hospital | Medical OOP: Other | 39 | 39.00 | 0.35 | 1.01 | 3.03 | 4.79 | 17.66 | 3.78 |
| Secondary hospital - General referral hospital | Medical OOP: Other purchases | 5 | 5.00 | 1.01 | 6.05 | 30.27 | 75.68 | 343.09 | 69.63 |
| Secondary hospital - General referral hospital | Non-Medical OOP | 161 | 146.00 | 0.25 | 5.87 | 10.09 | 17.44 | 160.95 | 11.58 |
| Secondary hospital - General referral hospital | Non-Medical OOP: Accommodation | 14 | 14.00 | 2.52 | 5.11 | 7.06 | 9.84 | 15.14 | 4.73 |
| Secondary hospital - General referral hospital | Non-Medical OOP: Food | 134 | 134.00 | 0.25 | 5.05 | 9.08 | 12.61 | 100.91 | 7.57 |
| Secondary hospital - General referral hospital | Non-Medical OOP: Other | 22 | 22.00 | 0.25 | 0.76 | 1.14 | 4.67 | 151.36 | 3.91 |
| Secondary hospital - General referral hospital | Non-Medical OOP: Transport | 66 | 66.00 | 0.25 | 1.58 | 3.78 | 6.05 | 37.84 | 4.48 |
| Secondary hospital - General referral hospital | Total OOP | 161 | 156.00 | 0.50 | 17.02 | 33.68 | 53.73 | 409.18 | 36.72 |
| Secondary hospital - General referral hospital | Total OOP excluding food related costs | 161 | 149.00 | 0.76 | 12.87 | 24.22 | 42.13 | 383.96 | 29.26 |
| Traditional practitioners | Medical OOP | 11 | 11.00 | 1.77 | 19.42 | 22.70 | 29.01 | 35.32 | 9.59 |
| Traditional practitioners | Medical OOP: Consumables | 5 | 5.00 | 2.52 | 3.03 | 3.18 | 5.05 | 18.16 | 2.02 |
| Traditional practitioners | Medical OOP: Medication | 2 | 2.00 | 17.66 | 19.55 | 21.44 | 23.34 | 25.23 | 3.78 |
| Traditional practitioners | Medical OOP: Other | 8 | 8.00 | 1.77 | 17.41 | 23.46 | 25.86 | 35.32 | 8.45 |
| Traditional practitioners | Non-Medical OOP | 11 | 8.00 | 1.51 | 2.84 | 4.41 | 7.69 | 15.64 | 4.86 |
| Traditional practitioners | Non-Medical OOP: Food | 8 | 8.00 | 1.26 | 1.51 | 3.41 | 7.69 | 8.58 | 6.18 |
| Traditional practitioners | Non-Medical OOP: Transport | 3 | 3.00 | 1.51 | 2.02 | 2.52 | 4.79 | 7.06 | 2.77 |
| Traditional practitioners | Total OOP | 11 | 11.00 | 10.85 | 20.18 | 27.75 | 31.61 | 37.84 | 11.43 |

| | | | | | | | | | |
|---|---|---|---|---|---|---|---|---|---|
| Traditional practitioners | Total OOP excluding food related costs | 11 | 11.00 | 8.83 | 19.42 | 23.21 | 29.01 | 35.32 | 9.59 |

**Supporting Information 16: Table Median OOP per screening strategy per screening outcome**

| ScreeningStrat | Outcome_PLNTHA | variable | n_total | n_nonzero | Min_Value | Q1 | Median | Q3 | Max_Value | IQR |
|---|---|---|---|---|---|---|---|---|---|---|
| Active | Serology: negative | Medical OOP: Consumables | 128 | 52 | 1.26 | 4.04 | 8.20 | 20.81 | 89.81 | 16.78 |
| Active | Serology: result unknown | Medical OOP: Consumables | 10 | 2 | 3.53 | 6.24 | 8.96 | 11.67 | 14.38 | 5.42 |
| Active | Serology: positive; Microscopy: not done/unknown | Medical OOP: Consumables | 13 | 2 | 2.52 | 4.41 | 6.31 | 8.20 | 10.09 | 3.78 |
| Active | Serology performed; Microscopy negative | Medical OOP: Consumables | 61 | 15 | 0.50 | 2.27 | 3.03 | 5.55 | 16.40 | 3.28 |
| Active | Confirmed HAT case | Medical OOP: Consumables | 18 | 6 | 0.76 | 0.95 | 1.51 | 2.27 | 5.55 | 1.32 |
| Passive | Serology: negative | Medical OOP: Consumables | 134 | 104 | 0.50 | 1.70 | 6.56 | 10.60 | 62.06 | 8.89 |
| Passive | Serology: result unknown | Medical OOP: Consumables | 11 | 8 | 1.01 | 1.83 | 4.54 | 11.23 | 17.66 | 9.40 |
| Passive | Serology: positive; Microscopy: not done/unknown | Medical OOP: Consumables | 9 | 6 | 0.76 | 2.77 | 15.14 | 25.23 | 27.75 | 22.45 |
| Passive | Serology performed; Microscopy negative | Medical OOP: Consumables | 32 | 20 | 0.50 | 0.76 | 1.39 | 11.35 | 63.07 | 10.60 |
| Passive | Confirmed HAT case | Medical OOP: Consumables | 28 | 23 | 0.50 | 1.51 | 3.53 | 11.35 | 104.94 | 9.84 |
| Active | Serology: negative | Medical OOP: Laboratory | 128 | 32 | 0.25 | 0.95 | 2.02 | 4.04 | 20.18 | 3.09 |
| Active | Serology: result unknown | Medical OOP: Laboratory | 10 | 1 | 3.03 | 3.03 | 3.03 | 3.03 | 3.03 | - |
| Active | Serology: positive; Microscopy: not done/unknown | Medical OOP: Laboratory | 13 | 1 | 1.01 | 1.01 | 1.01 | 1.01 | 1.01 | - |
| Active | Serology performed; Microscopy negative | Medical OOP: Laboratory | 61 | 9 | 0.50 | 0.50 | 0.50 | 2.52 | 4.04 | 2.02 |
| Active | Confirmed HAT case | Medical OOP: Laboratory | 18 | 2 | 4.04 | 4.54 | 5.05 | 5.55 | 6.05 | 1.01 |
| Passive | Serology: negative | Medical OOP: Laboratory | 134 | 32 | 0.25 | 1.20 | 1.64 | 2.52 | 28.25 | 1.32 |
| Passive | Serology: result unknown | Medical OOP: Laboratory | 11 | 3 | 0.76 | 1.01 | 1.26 | 2.27 | 3.28 | 1.26 |
| Passive | Serology: positive; Microscopy: not done/unknown | Medical OOP: Laboratory | 9 | 2 | 0.10 | 0.58 | 1.06 | 1.54 | 2.02 | 0.96 |
| Passive | Serology performed; Microscopy negative | Medical OOP: Laboratory | 32 | 8 | 0.76 | 1.01 | 1.51 | 1.77 | 3.53 | 0.76 |
| Passive | Confirmed HAT case | Medical OOP: Laboratory | 28 | 11 | 0.25 | 0.50 | 1.77 | 4.54 | 7.57 | 4.04 |
| Active | Serology: negative | Medical OOP: Other purchases | 128 | 4 | 1.01 | 9.71 | 21.44 | 108.48 | 343.09 | 98.76 |
| Active | Serology: result unknown | Medical OOP: Other purchases | 10 | 0 | | | | | | |
| Active | Serology: positive; Microscopy: not done/unknown | Medical OOP: Other purchases | 13 | 0 | | | | | | |
| Active | Serology performed; Microscopy negative | Medical OOP: Other purchases | 61 | 0 | | | | | | |
| Active | Confirmed HAT case | Medical OOP: Other purchases | 18 | 1 | 6.05 | 6.05 | 6.05 | 6.05 | 6.05 | - |
| Passive | Serology: negative | Medical OOP: Other purchases | 134 | 1 | 75.68 | 75.68 | 75.68 | 75.68 | 75.68 | - |
| Passive | Serology: result unknown | Medical OOP: Other purchases | 11 | 0 | | | | | | |
| Passive | Serology: positive; Microscopy: not done/unknown | Medical OOP: Other purchases | 9 | 0 | | | | | | |
| Passive | Serology performed; Microscopy negative | Medical OOP: Other purchases | 32 | 0 | | | | | | |
| Passive | Confirmed HAT case | Medical OOP: Other purchases | 28 | 3 | 1.01 | 1.26 | 1.51 | 25.98 | 50.45 | 24.72 |
| Active | Serology: negative | Medical OOP: Medication | 128 | 50 | 0.25 | 1.83 | 8.20 | 21.70 | 74.77 | 19.87 |
| Active | Serology: result unknown | Medical OOP: Medication | 10 | 2 | 0.66 | 4.09 | 7.52 | 10.95 | 14.38 | 6.86 |
| Active | Serology: positive; Microscopy: not done/unknown | Medical OOP: Medication | 13 | 2 | 0.25 | 0.95 | 1.64 | 2.33 | 3.03 | 1.39 |
| Active | Serology performed; Microscopy negative | Medical OOP: Medication | 61 | 12 | 0.50 | 2.08 | 5.05 | 7.76 | 27.75 | 5.68 |
| Active | Confirmed HAT case | Medical OOP: Medication | 18 | 2 | 2.02 | 3.91 | 5.80 | 7.69 | 9.59 | 3.78 |
| Passive | Serology: negative | Medical OOP: Medication | 134 | 64 | 0.50 | 6.69 | 12.61 | 17.66 | 85.77 | 10.97 |
| Passive | Serology: result unknown | Medical OOP: Medication | 11 | 7 | 0.50 | 2.77 | 7.57 | 20.31 | 31.79 | 17.53 |
| Passive | Serology: positive; Microscopy: not done/unknown | Medical OOP: Medication | 9 | 4 | 3.43 | 4.64 | 6.56 | 11.73 | 22.70 | 7.09 |
| Passive | Serology performed; Microscopy negative | Medical OOP: Medication | 32 | 12 | 2.02 | 3.28 | 7.32 | 15.51 | 59.54 | 12.24 |
| Passive | Confirmed HAT case | Medical OOP: Medication | 28 | 17 | 1.01 | 2.52 | 8.98 | 46.92 | 121.09 | 44.40 |
| Active | Serology: negative | Medical OOP: Hospitalization | 128 | 10 | 2.52 | 9.71 | 11.60 | 13.37 | 19.17 | 3.66 |
| Active | Serology: result unknown | Medical OOP: Hospitalization | 10 | 0 | | | | | | |
| Active | Serology: positive; Microscopy: not done/unknown | Medical OOP: Hospitalization | 13 | 0 | | | | | | |

| | | | | | | | | | | |
|---|---|---|---|---|---|---|---|---|---|---|
| Active | Serology performed; Microscopy negative | Medical OOP: Hospitalization | 61 | 3 | 1.01 | 1.14 | 1.26 | 8.20 | 15.14 | 7.06 |
| Active | Confirmed HAT case | Medical OOP: Hospitalization | 18 | 3 | 6.31 | 6.69 | 7.06 | 8.58 | 10.09 | 1.89 |
| Passive | Serology: negative | Medical OOP: Hospitalization | 134 | 19 | 2.52 | 3.53 | 4.29 | 6.81 | 15.64 | 3.28 |
| Passive | Serology: result unknown | Medical OOP: Hospitalization | 11 | 1 | 3.53 | 3.53 | 3.53 | 3.53 | 3.53 | - |
| Passive | Serology: positive; Microscopy: not done/unknown | Medical OOP: Hospitalization | 9 | 3 | 1.26 | 3.66 | 6.05 | 9.59 | 13.12 | 5.93 |
| Passive | Serology performed; Microscopy negative | Medical OOP: Hospitalization | 32 | 2 | 4.54 | 5.93 | 7.32 | 8.70 | 10.09 | 2.77 |
| Passive | Confirmed HAT case | Medical OOP: Hospitalization | 28 | 9 | 2.52 | 6.05 | 7.57 | 17.66 | 30.27 | 11.60 |
| Active | Serology: negative | Medical OOP: Other | 128 | 10 | 0.10 | 0.28 | 0.50 | 2.40 | 50.45 | 2.12 |
| Active | Serology: result unknown | Medical OOP: Other | 10 | 0 | | | | | | |
| Active | Serology: positive; Microscopy: not done/unknown | Medical OOP: Other | 13 | 1 | 0.35 | 0.35 | 0.35 | 0.35 | 0.35 | - |
| Active | Serology performed; Microscopy negative | Medical OOP: Other | 61 | 1 | 0.50 | 0.50 | 0.50 | 0.50 | 0.50 | - |
| Active | Confirmed HAT case | Medical OOP: Other | 18 | 0 | | | | | | |
| Passive | Serology: negative | Medical OOP: Other | 134 | 30 | 0.50 | 1.51 | 2.77 | 4.41 | 17.66 | 2.90 |
| Passive | Serology: result unknown | Medical OOP: Other | 11 | 2 | 1.01 | 1.39 | 1.77 | 2.14 | 2.52 | 0.76 |
| Passive | Serology: positive; Microscopy: not done/unknown | Medical OOP: Other | 9 | 1 | 25.23 | 25.23 | 25.23 | 25.23 | 25.23 | - |
| Passive | Serology performed; Microscopy negative | Medical OOP: Other | 32 | 3 | 5.05 | 7.32 | 9.59 | 16.15 | 22.70 | 8.83 |
| Passive | Confirmed HAT case | Medical OOP: Other | 28 | 6 | 10.09 | 16.90 | 22.45 | 26.87 | 35.32 | 9.96 |
| Active | Serology: negative | Non-Medical OOP: Transport | 128 | 25 | 0.50 | 2.88 | 5.80 | 9.59 | 37.84 | 6.71 |
| Active | Serology: result unknown | Non-Medical OOP: Transport | 10 | 1 | 1.01 | 1.01 | 1.01 | 1.01 | 1.01 | - |
| Active | Serology: positive; Microscopy: not done/unknown | Non-Medical OOP: Transport | 13 | 0 | | | | | | |
| Active | Serology performed; Microscopy negative | Non-Medical OOP: Transport | 61 | 4 | 0.76 | 0.76 | 1.26 | 1.89 | 2.27 | 1.14 |
| Active | Confirmed HAT case | Non-Medical OOP: Transport | 18 | 5 | 0.20 | 2.52 | 3.03 | 5.05 | 6.56 | 2.52 |
| Passive | Serology: negative | Non-Medical OOP: Transport | 134 | 43 | 0.25 | 1.01 | 1.51 | 5.05 | 17.15 | 4.04 |
| Passive | Serology: result unknown | Non-Medical OOP: Transport | 11 | 5 | 1.01 | 1.51 | 3.03 | 3.53 | 5.05 | 2.02 |
| Passive | Serology: positive; Microscopy: not done/unknown | Non-Medical OOP: Transport | 9 | 1 | 5.05 | 5.05 | 5.05 | 5.05 | 5.05 | - |
| Passive | Serology performed; Microscopy negative | Non-Medical OOP: Transport | 32 | 6 | 0.50 | 2.40 | 3.78 | 4.79 | 7.57 | 2.40 |
| Passive | Confirmed HAT case | Non-Medical OOP: Transport | 28 | 12 | 0.50 | 1.70 | 5.80 | 25.67 | 51.97 | 23.97 |
| Active | Serology: negative | Non-Medical OOP: Food | 128 | 50 | 0.10 | 1.51 | 5.42 | 10.09 | 35.82 | 8.58 |
| Active | Serology: result unknown | Non-Medical OOP: Food | 10 | 1 | 13.62 | 13.62 | 13.62 | 13.62 | 13.62 | - |
| Active | Serology: positive; Microscopy: not done/unknown | Non-Medical OOP: Food | 13 | 0 | | | | | | |
| Active | Serology performed; Microscopy negative | Non-Medical OOP: Food | 61 | 10 | 0.10 | 0.48 | 2.40 | 5.49 | 12.61 | 5.01 |
| Active | Confirmed HAT case | Non-Medical OOP: Food | 18 | 9 | 1.01 | 5.05 | 10.09 | 12.11 | 32.80 | 7.06 |
| Passive | Serology: negative | Non-Medical OOP: Food | 134 | 92 | 0.10 | 1.01 | 4.29 | 10.22 | 70.64 | 9.21 |
| Passive | Serology: result unknown | Non-Medical OOP: Food | 11 | 6 | 4.04 | 5.05 | 7.06 | 11.73 | 16.65 | 6.69 |
| Passive | Serology: positive; Microscopy: not done/unknown | Non-Medical OOP: Food | 9 | 5 | 15.14 | 16.65 | 25.23 | 30.27 | 63.07 | 13.62 |
| Passive | Serology performed; Microscopy negative | Non-Medical OOP: Food | 32 | 15 | 0.50 | 3.78 | 10.60 | 12.61 | 20.18 | 8.83 |
| Passive | Confirmed HAT case | Non-Medical OOP: Food | 28 | 24 | 1.51 | 6.62 | 9.96 | 17.91 | 151.36 | 11.29 |
| Active | Serology: negative | Non-Medical OOP: Accommodation | 128 | 9 | 1.01 | 3.53 | 7.06 | 15.14 | 17.66 | 11.60 |
| Active | Serology: result unknown | Non-Medical OOP: Accommodation | 10 | 0 | | | | | | |
| Active | Serology: positive; Microscopy: not done/unknown | Non-Medical OOP: Accommodation | 13 | 0 | | | | | | |
| Active | Serology performed; Microscopy negative | Non-Medical OOP: Accommodation | 61 | 0 | | | | | | |
| Active | Confirmed HAT case | Non-Medical OOP: Accommodation | 18 | 0 | | | | | | |
| Passive | Serology: negative | Non-Medical OOP: Accommodation | 134 | 4 | 5.30 | 6.62 | 8.83 | 11.73 | 15.14 | 5.11 |
| Passive | Serology: result unknown | Non-Medical OOP: Accommodation | 11 | 1 | 4.29 | 4.29 | 4.29 | 4.29 | 4.29 | - |
| Passive | Serology: positive; Microscopy: not done/unknown | Non-Medical OOP: Accommodation | 9 | 0 | | | | | | |
| Passive | Serology performed; Microscopy negative | Non-Medical OOP: Accommodation | 32 | 0 | | | | | | |

| | | | | | | | | | | |
|---|---|---|---|---|---|---|---|---|---|---|
| Passive | Confirmed HAT case | Non-Medical OOP: Accommodation | 28 | 0 | | | | | | |
| Active | Serology: negative | Non-Medical OOP: Other | 128 | 13 | 0.25 | 0.50 | 0.76 | 1.01 | 9.59 | 0.50 |
| Active | Serology: result unknown | Non-Medical OOP: Other | 10 | 0 | | | | | | |
| Active | Serology: positive; Microscopy: not done/unknown | Non-Medical OOP: Other | 13 | 0 | | | | | | |
| Active | Serology performed; Microscopy negative | Non-Medical OOP: Other | 61 | 2 | 0.25 | 0.50 | 0.76 | 1.01 | 1.26 | 0.50 |
| Active | Confirmed HAT case | Non-Medical OOP: Other | 18 | 1 | 0.50 | 0.50 | 0.50 | 0.50 | 0.50 | - |
| Passive | Serology: negative | Non-Medical OOP: Other | 134 | 19 | 0.25 | 0.25 | 0.50 | 1.79 | 151.36 | 1.54 |
| Passive | Serology: result unknown | Non-Medical OOP: Other | 11 | 0 | | | | | | |
| Passive | Serology: positive; Microscopy: not done/unknown | Non-Medical OOP: Other | 9 | 1 | 30.27 | 30.27 | 30.27 | 30.27 | 30.27 | - |
| Passive | Serology performed; Microscopy negative | Non-Medical OOP: Other | 32 | 4 | 3.53 | 6.18 | 9.21 | 17.60 | 36.33 | 11.42 |
| Passive | Confirmed HAT case | Non-Medical OOP: Other | 28 | 5 | 0.50 | 1.01 | 2.52 | 7.57 | 10.09 | 6.56 |
| Active | Serology: negative | Medical OOP | 128 | 57 | 0.25 | 6.05 | 20.18 | 72.40 | 376.14 | 66.35 |
| Active | Serology: result unknown | Medical OOP | 10 | 3 | 0.66 | 2.09 | 3.53 | 17.66 | 31.79 | 15.57 |
| Active | Serology: positive; Microscopy: not done/unknown | Medical OOP | 13 | 3 | 1.26 | 1.89 | 2.52 | 8.00 | 13.47 | 6.10 |
| Active | Serology performed; Microscopy negative | Medical OOP | 61 | 18 | 0.50 | 1.96 | 7.04 | 11.16 | 44.15 | 9.21 |
| Active | Confirmed HAT case | Medical OOP | 18 | 9 | 1.51 | 2.52 | 5.55 | 10.09 | 19.93 | 7.57 |
| Passive | Serology: negative | Medical OOP | 134 | 110 | 0.25 | 6.81 | 17.15 | 26.87 | 242.68 | 20.06 |
| Passive | Serology: result unknown | Medical OOP | 11 | 9 | 1.51 | 5.80 | 18.16 | 28.76 | 34.31 | 22.96 |
| Passive | Serology: positive; Microscopy: not done/unknown | Medical OOP | 9 | 7 | 5.05 | 5.25 | 8.93 | 41.25 | 66.09 | 36.00 |
| Passive | Serology performed; Microscopy negative | Medical OOP | 32 | 20 | 0.76 | 4.29 | 12.87 | 31.60 | 72.65 | 27.31 |
| Passive | Confirmed HAT case | Medical OOP | 28 | 26 | 1.51 | 6.94 | 20.69 | 66.21 | 196.67 | 59.27 |
| Active | Serology: negative | Non-Medical OOP | 128 | 54 | 0.10 | 1.77 | 6.31 | 22.33 | 88.55 | 20.56 |
| Active | Serology: result unknown | Non-Medical OOP | 10 | 1 | 14.63 | 14.63 | 14.63 | 14.63 | 14.63 | - |
| Active | Serology: positive; Microscopy: not done/unknown | Non-Medical OOP | 13 | 0 | | | | | | |
| Active | Serology performed; Microscopy negative | Non-Medical OOP | 61 | 11 | 0.25 | 0.81 | 1.77 | 5.80 | 13.87 | 4.99 |
| Active | Confirmed HAT case | Non-Medical OOP | 18 | 12 | 0.20 | 4.41 | 7.57 | 12.49 | 32.80 | 8.07 |
| Passive | Serology: negative | Non-Medical OOP | 134 | 106 | 0.10 | 1.15 | 5.17 | 12.61 | 160.95 | 11.47 |
| Passive | Serology: result unknown | Non-Medical OOP | 11 | 7 | 1.01 | 6.05 | 9.08 | 12.11 | 24.47 | 6.05 |
| Passive | Serology: positive; Microscopy: not done/unknown | Non-Medical OOP | 9 | 5 | 16.65 | 20.18 | 25.23 | 30.27 | 93.34 | 10.09 |
| Passive | Serology performed; Microscopy negative | Non-Medical OOP | 32 | 17 | 0.50 | 5.55 | 11.10 | 17.66 | 51.21 | 12.11 |
| Passive | Confirmed HAT case | Non-Medical OOP | 28 | 26 | 3.53 | 6.24 | 10.09 | 17.05 | 161.45 | 10.81 |
| Active | Serology: negative | Total OOP | 128 | 63 | 0.10 | 7.64 | 18.52 | 73.97 | 409.18 | 66.32 |
| Active | Serology: result unknown | Total OOP | 10 | 3 | 0.66 | 2.09 | 3.53 | 24.97 | 46.42 | 22.88 |
| Active | Serology: positive; Microscopy: not done/unknown | Total OOP | 13 | 3 | 1.26 | 1.89 | 2.52 | 8.00 | 13.47 | 6.10 |
| Active | Serology performed; Microscopy negative | Total OOP | 61 | 21 | 0.25 | 1.36 | 3.53 | 12.61 | 49.70 | 11.25 |
| Active | Confirmed HAT case | Total OOP | 18 | 14 | 0.20 | 5.17 | 9.84 | 19.49 | 37.59 | 14.32 |
| Passive | Serology: negative | Total OOP | 134 | 119 | 0.25 | 7.44 | 20.18 | 36.45 | 263.37 | 29.01 |
| Passive | Serology: result unknown | Total OOP | 11 | 11 | 1.01 | 6.31 | 18.16 | 31.66 | 58.02 | 25.35 |
| Passive | Serology: positive; Microscopy: not done/unknown | Total OOP | 9 | 7 | 5.45 | 22.12 | 51.46 | 79.59 | 98.39 | 57.47 |
| Passive | Serology performed; Microscopy negative | Total OOP | 32 | 21 | 0.76 | 7.06 | 24.72 | 47.43 | 88.29 | 40.36 |
| Passive | Confirmed HAT case | Total OOP | 28 | 28 | 6.56 | 12.49 | 24.52 | 81.71 | 358.12 | 69.22 |
| Active | Serology: negative | Total OOP excl Food | 128 | 61.00 | 0.25 | 6.05 | 15.14 | 59.54 | 383.96 | 53.48 |
| Active | Serology: result unknown | Total OOP excl Food | 10 | 3.00 | 0.66 | 2.09 | 3.53 | 18.16 | 32.80 | 16.07 |
| Active | Serology: positive; Microscopy: not done/unknown | Total OOP excl Food | 13 | 3.00 | 1.26 | 1.89 | 2.52 | 8.00 | 13.47 | 6.10 |
| Active | Serology performed; Microscopy negative | Total OOP excl Food | 61 | 18.00 | 0.50 | 1.96 | 7.04 | 12.30 | 45.91 | 10.34 |
| Active | Confirmed HAT case | Total OOP excl Food | 18 | 11.00 | 0.20 | 3.53 | 5.05 | 10.85 | 22.96 | 7.32 |

| | | | | | | | | | | |
|---|---|---|---|---|---|---|---|---|---|---|
| Passive | Serology: negative | Total OOP excl Food | 134 | 112.00 | 0.76 | 7.00 | 17.53 | 29.45 | 250.76 | 22.45 |
| Passive | Serology: result unknown | Total OOP excl Food | 11 | 11.00 | 1.01 | 3.78 | 12.87 | 24.47 | 41.37 | 20.69 |
| Passive | Serology: positive; Microscopy: not done/unknown | Total OOP excl Food | 9 | 7.00 | 5.05 | 7.19 | 35.32 | 43.77 | 66.09 | 36.58 |
| Passive | Serology performed; Microscopy negative | Total OOP excl Food | 32 | 21.00 | 0.76 | 4.79 | 14.13 | 29.77 | 83.75 | 24.97 |
| Passive | Confirmed HAT case | Total OOP excl Food | 28 | 26.00 | 2.02 | 7.69 | 22.83 | 66.21 | 206.76 | 58.51 |
| Active | Serology: negative | Nb_struct | 128 | 128 | 1.00 | 1.00 | 1.00 | 3.00 | 6.00 | 2.00 |
| Active | Serology: result unknown | Nb_struct | 10 | 10 | 1.00 | 1.00 | 1.50 | 2.00 | 5.00 | 1.00 |
| Active | Serology: positive; Microscopy: not done/unknown | Nb_struct | 13 | 13 | 1.00 | 1.00 | 1.00 | 2.00 | 4.00 | 1.00 |
| Active | Serology performed; Microscopy negative | Nb_struct | 61 | 61 | 1.00 | 1.00 | 1.00 | 2.00 | 4.00 | 1.00 |
| Active | Confirmed HAT case | Nb_struct | 18 | 18 | 1.00 | 1.00 | 2.00 | 2.00 | 5.00 | 1.00 |
| Passive | Serology: negative | Nb_struct | 134 | 134 | 1.00 | 1.00 | 1.00 | 2.00 | 8.00 | 1.00 |
| Passive | Serology: result unknown | Nb_struct | 11 | 11 | 1.00 | 1.00 | 2.00 | 2.00 | 2.00 | 1.00 |
| Passive | Serology: positive; Microscopy: not done/unknown | Nb_struct | 9 | 9 | 1.00 | 1.00 | 1.00 | 3.00 | 3.00 | 2.00 |
| Passive | Serology performed; Microscopy negative | Nb_struct | 32 | 32 | 1.00 | 1.00 | 1.00 | 1.00 | 3.00 | - |
| Passive | Confirmed HAT case | Nb_struct | 28 | 28 | 1.00 | 1.00 | 2.00 | 3.00 | 4.00 | 2.00 |

**Supporting Information 17: Frequency of the structures visited by participants**

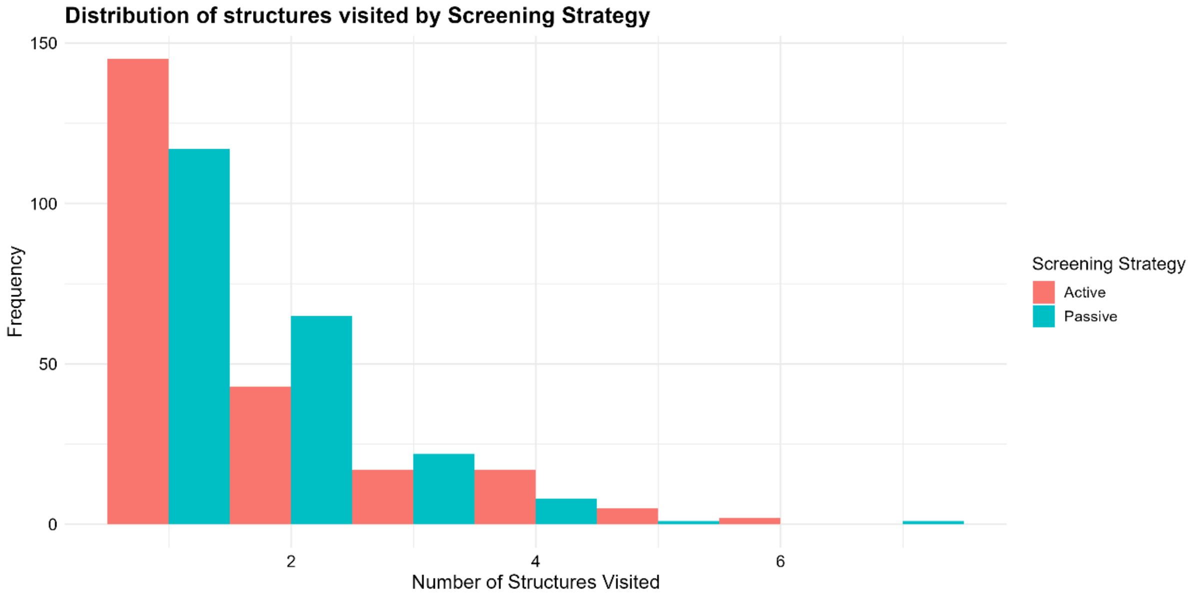

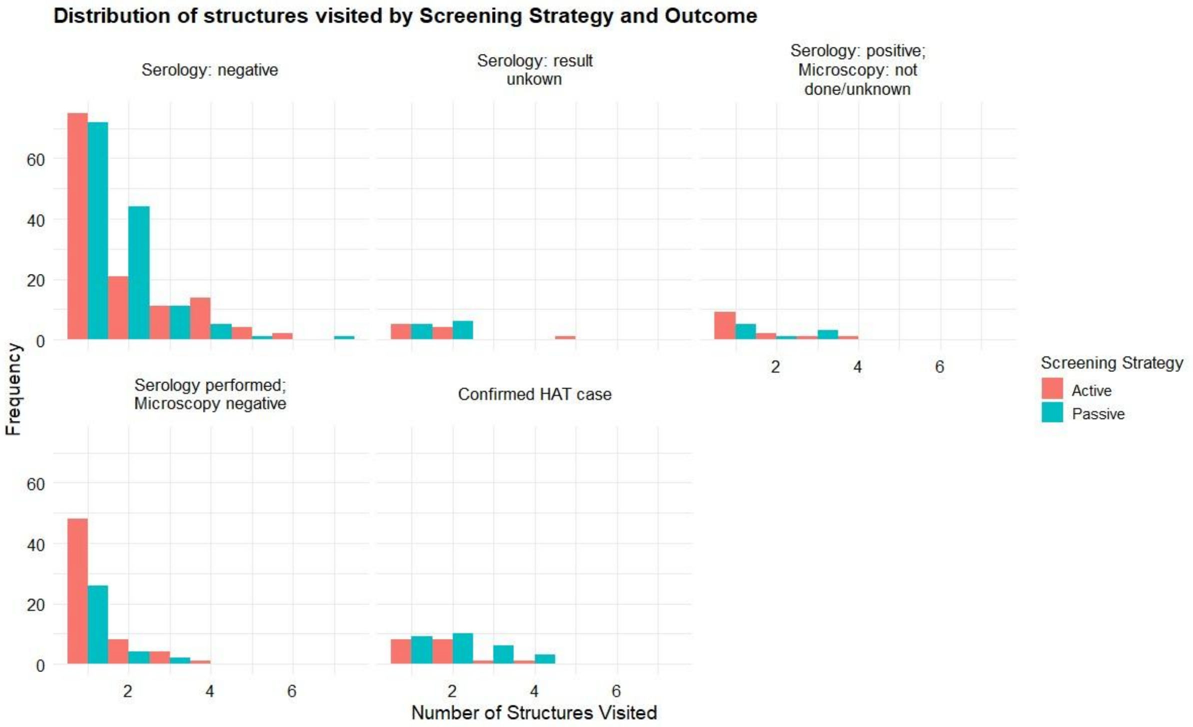

**Supporting Information 18: Interview guide – Individual semi-structured interviews**

For **Supporting Information 18: Interview guide – Individual semi-structured interviews**, please contact the first author, as this guide was developed in French and medRxiv does not permit supplementary information to be submitted in languages other than English.

**Supporting Information 19: Interview guide – Focus group discussions**

For **Supporting Information 19: Interview guide – Focus group discussions**, please contact the first author, as this guide was developed in French and medRxiv does not permit supplementary information to be submitted in languages other than English.

**Supporting Information 20: Survey « Evaluation des dépenses médicales et non-médicales THA »**

For **Supporting Information 20: Survey**, please contact the first author, as this guide was developed in French and medRxiv does not permit supplementary information to be submitted in languages other than English.
